# Dynamics of gut mucosal colonisation with extended spectrum beta-lactamase producing Enterobacterales in Malawi

**DOI:** 10.1101/2021.10.08.21264775

**Authors:** Joseph M. Lewis, Madalitso Mphasa, Rachel Banda, Mathew A. Beale, Eva Heinz, Jane Mallewa, Christopher Jewell, Brian Faragher, Nicholas R. Thomson, Nicholas A Feasey

## Abstract

Shortening courses of antimicrobials has been proposed to reduce risk of antimicrobial resistant (AMR) infections, but acquisition and selection dynamics under antimicrobial pressure at the individual level are poorly understood. We combine multi-state modelling and whole-genome sequencing to understand colonisation dynamics of extended-spectrum beta-lactamase producing Enterobacterales (ESBL-E) in Malawian adults. We demonstrate prolonged post-exposure antibiotic effect, meaning short courses exert similar colonisation pressure to longer ones. Genome data does not identify widespread hospital-associated ESBL-E transmission, hence apparent acquisitions may be selected from the patient microbiota by antimicrobial exposure. Understanding ESBL-E dynamics under antimicrobial pressure is crucial for evidence-based stewardship protocols.

## Introduction

Antimicrobials are one of the most successful therapies available to modern medicine, but the spread of antimicrobial resistance (AMR) is a threat to their effective use. Significant global effort is being directed at antimicrobial stewardship programmes designed to optimise antimicrobial use, both to avoid dispensing these agents where not warranted and, where they are deemed to be necessary, minimising both duration of exposure and spectrum of bacteria affected^1^. These principles are guided in part by well-described population level associations between antimicrobial exposure and prevalence of AMR at multiple spatial and temporal scales^2–4^. Antimicrobial stewardship interventions at the individual level often emphasise rationalisation of antimicrobials through narrowing their spectrum of action as soon as possible after commencement of broad empiric antimicrobial therapy in severely unwell individuals. The time frame (e.g., 48 hours) for this is typically pragmatically selected based on likely availability of diagnostic test results. This rationalisation of therapy is, in part, based on the assumption that it will reduce emergence of AMR but at the individual patient level, the mechanism by which antimicrobial exposure acts to promote colonisation and/or infection with resistant pathogens, and the dynamics of colonisation and decolonisation, are not well understood^5–8^. Improved understanding of the dynamics of individual-level AMR-acquisition under antimicrobial pressure is therefore necessary as it has the potential to optimise stewardship protocols.

One setting in which antimicrobial stewardship is a significant challenge is in the treatment of severe febrile illness in the low- and middle-income countries (LMIC) of sub-Saharan Africa (sSA). In Blantyre, Malawi, for example, as in much of sSA, limited availability of diagnostics results in prolonged courses of broad-spectrum antimicrobials – largely ceftriaxone, a third-generation cephalosporin (3GC) antibiotic^9^ -for people hospitalised with severe febrile illness. Ceftriaxone has a favourable spectrum and pharmacokinetics for locally circulating bacterial pathogens and hence has been extensively used since its introduction to the Malawian national formulary in 2005^10^. However, this widespread use has been associated with an increase in 3GC resistance, particularly in bacteria of the order Enterobacterales, and largely mediated by extended spectrum beta-lactamase (ESBL) enzymes^11^. ESBL-producing Enterobacterales (henceforth ESBL-E) are an increasing public health challenge throughout much of sSA^12,13^, and often have few or no locally available treatment options; in Blantyre, 91% of invasive *K. pneumoniae* are now ESBL producers^11^, and strategies to reduce ESBL-E infections are needed.

In ESBL-E, gut mucosal colonisation is thought to precede invasive infection.^14–16^ It is common across sSA, and has often been found to be associated with prior hospitalization and/or antimicrobial exposure^13^. An improved mechanistic understanding of colonisation dynamics following these exposures therefore has the potential to inform evidence-based interventions to reduce colonisation and hence opportunity for transmission. Here we present the results from a clinical study of longitudinal ESBL-E carriage in Blantyre, Malawi, sampling adults as they pass through the hospital and are exposed to antimicrobials. We use multi-state modelling^17^ and whole-genome sequencing as a high-resolution bacterial typing tool to describe and understand the dynamics of ESBL-E colonisation and de-colonisation.

## Results

### Antimicrobial exposure is associated with rapid and prolonged increase in ESBL-E prevalence

Between 19^th^ February 2017 and 2^nd^ October 2018, we recruited 425 adults; i) 225 patients with sepsis and antimicrobial exposure, admitted to Queen Elizabeth Central hospital (QECH) Blantyre; ii) 100 antimicrobial-unexposed inpatients and iii) 100 antimicrobial-unexposed community members (Table 1, Supplementary Figure 1). Participants were typically young (median age 35.6 years), 73/425 (41%) were HIV infected with a high proportion of these (144/173, 83%) established on antiretroviral therapy (ART) and lifelong co-trimoxazole preventative therapy (CPT, 110/171, 64%) as per WHO guidance. 1417 stool samples in total were collected from the participants, and one or more ESBL-E species was cultured in 723/1417 (51%) of samples. 1032 organisms were isolated, most commonly *E. coli* (n = 686) followed by members of the *K. pneumoniae* species complex (n = 245, Supplementary Table 1 and Supplementary Figures 2 and 3).

**Table 1:** Baseline characteristics of included participants

| Variable | Sepsis (n=225) | Inpatient (n=100) | Community (n=100) | p | Total (n = 425) |
| --- | --- | --- | --- | --- | --- |
| **Demographics** |  |  |  |  |  |
| Age (yrs) | 35.9 (27.8-43.5) | 40.4 (29.1-48.3) | 32.5 (24.0-38.4) | <0.001 | 35.6 (26.9-43.9) |
| Male | 114/225 (51%) | 51/100 (51%) | 40/100 (40%) | 0.163 | 205/425 (48%) |
| **HIV status** |  |  |  |  |  |
| HIV Infected | 143/225 (64%) | 12/100 (12%) | 18/100 (18%) | <0.001 | 173/425 (41%) |
| HIV Uninfected | 70/225 (31%) | 77/100 (77%) | 22/100 (22%) |  | 169/425 (40%) |
| HIV Unknown | 12/225 (5%) | 11/100 (11%) | 60/100 (60%) |  | 83/425 (20%) |
| **ART status\*** |  |  |  |  |  |
| Current CPT | 98/141 (70%) | 5/12 (42%) | 7/18 (39%) | 0.013 | 110/171 (64%) |
| Current ART | 117/143 (82%) | 9/12 (75%) | 18/18 (100%) | 0.082 | 144/173 (83%) |
| Months on ART | 28.7 (3.7-72.6) | 35.1 (2.9-79.8) | 31.5 (13.0-79.9) | 0.698 | 29.5 (3.8-72.8) |
| **Healthcare exposure** |  |  |  |  |  |
| Antibiotics within 28 days† | 60/225 (27%) | 0/100 (0%) | 0/100 (0%) | <0.001 | 60/425 (14%) |
| Hospitalised within 28 days | 18/225 (8%) | 1/100 (1%) | 0/100 (0%) | <0.001 | 19/425 (4%) |
| Current TB treatment | 10/225 (4%) | 0/100 (0%) | 4/100 (4%) | 0.083 | 14/425 (3%) |
| **Household** |  |  |  |  |  |
| Number of adults | 2.0 (2.0-3.0) | 3.0 (2.0-4.0) | 2.0 (2.0-4.0) | 0.907 | 3.0 (2.0-4.0) |
| Number of children | 2.0 (1.0-3.0) | 2.0 (1.0-3.0) | 2.0 (1.0-3.0) | 0.395 | 2.0 (1.0-3.0) |
| Keep animals | 71/225 (32%) | 43/100 (43%) | 15/100 (15%) | <0.001 | 129/425 (30%) |
| Poultry | 46/71 (65%) | 34/43 (79%) | 10/15 (67%) |  | 90/129 (70%) |
| Dogs | 18/71 (25%) | 11/43 (26%) | 9/15 (60%) |  | 38/129 (29%) |
| Goats | 12/71 (17%) | 7/43 (16%) | 1/15 (7%) |  | 20/129 (16%) |
| Other | 3/71 (4%) | 6/43 (14%) | 0/15 (0%) |  | 9/129 (7%) |
| Electricity in house | 119/225 (53%) | 41/100 (41%) | 58/100 (58%) | 0.041 | 218/425 (51%) |
| Flush toilet‡ | 14/225 (6%) | 5/100 (5%) | 1/100 (1%) | 0.110 | 20/425 (5%) |
| Protected water source§ | 216/225 (96%) | 92/100 (92%) | 98/100 (98%) | 0.124 | 406/425 (96%) |
| Treat drinking water with chlorine | 19/225 (8%) | 5/100 (5%) | 0/100 (0%) | 0.004 | 24/425 (6%) |

ART = Antiretroviral therapy, CPT = Co-trimoxazole preventative therapy, TB = tuberculosis. Numeric variables are presented as median (IQR) and categorical variables as proportions. P-values are from Fisher's exact test or Kruskal-Wallis tests (categorical or continuous variables respectively) across the three groups; p-value for HIV status compares distribution of HIV status across the three groups. In some cases denominator may be less than the total number of participants due to missing data.
\* Denominator for ART status is HIV reactive participants only † Excluding TB treatment and CPT ‡ Flush toilet vs latrine (pit or hanging) or no toilet § Protected water source includes borehole, water piped into or outside dwelling or public standpipe; unprotected sources include surface water or unprotected springs.

**Figure 1:**
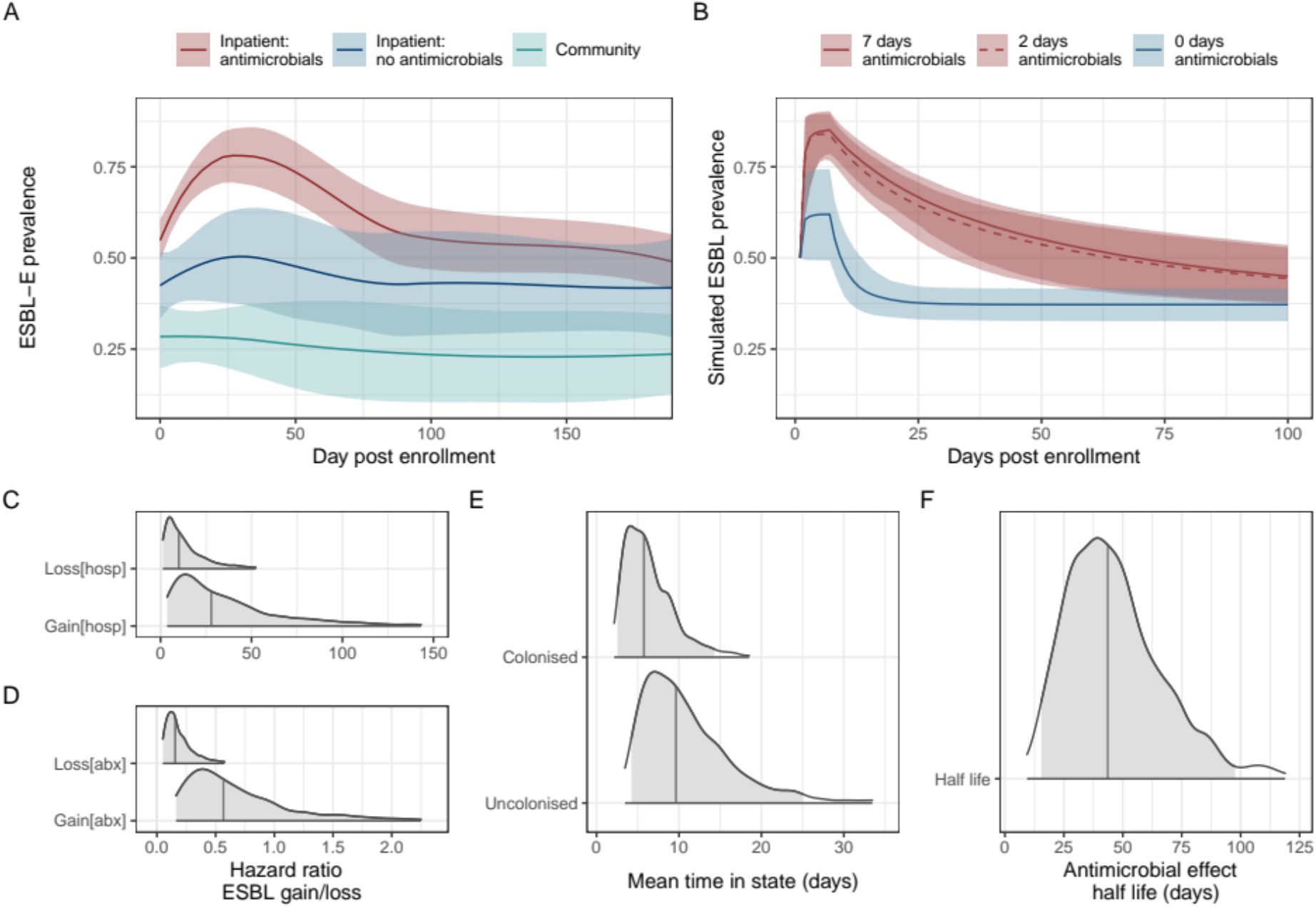
Prevalence and determinants of longitudinal ESBL-E carriage. **A:** ESBL prevalence stratified by the three study groups; inpatients exposed to antimicrobials (red), inpatients without antimicrobial exposure (blue), community members (green), showing sharp increase in prevalence following antimicrobial exposure. Prevalence is estimated using a LOESS non-parametric regression with 95% confidence interval. Community members are censored on antimicrobial exposure or hospitalization and antimicrobial-unexposed inpatients on antimicrobial exposure. **B:** simulated ESBL-E prevalence using final fitted model for a hypothetical cohort of patients with initial ESBL-E colonisation prevalence 50%, admitted to hospital for seven days and exposed to seven, two or zero days of antimicrobials, showing that there is little difference between seven and two days. **C-E:** posterior estimates of parameter values from final fitted model. Shaded grey areas shows 95% credible interval and vertical line shows median parameter estimate. **C:** hazard ratio of gain or loss of ESBL-E (expressed as natural logarithm) showing that antimicrobial exposure acts primarily to prolong carriage by reducing ESBL-E loss, whereas hospitalization acts to increase both gain and loss, with a net effect to increase prevalence. **D:** Mean time in colonised/uncolonised states with all covariate values set to 0 (i.e. no hospitalization or antimicrobial exposure). **E:** Half-life of effect of antimicrobial exposure, showing that antimicrobial exposure acts with a prolonged effect to prolong colonisation.

Baseline prevalence of ESBL-E colonization was 178/420 42% (95% CI 38-47%). In univariable modelling, HIV infection was associated with baseline ESBL-E colonization, an effect that multivariable logistic regression modelling suggested was largely driven by CPT exposure (Table 2). Other multivariable associations of ESBL-E colonization at enrolment (unprotected drinking and other household water sources [aOR of colonization 2.96 95% CI 1.07-8.75], sample collection in rainy season [aOR 2.21 95% CI 1.07-8.75], number of adults in the household [aOR 1.20 95% CI 1.03-1.40] and recent hospitalization [aOR 6.64 95% CI 1.98 – 30.75]), suggest importance of faecal-oral and within-household ESBL-E community transmission routes, as well as healthcare exposure in driving ESBL-E colonisation. ESBL-E prevalence at baseline was higher in the participants recruited in hospital (Figure 1A and Supplementary Table 1) than community members, a finding that was explained by prior healthcare exposure and increased HIV prevalence (and hence CPT exposure) in the former group (Tables 1 and 2).

**Table 2:** Univariable and multivariable associations of baseline ESBL colonisation

| Variable | Univariable |  | Multivariable |  |
| --- | --- | --- | --- | --- |
|  | OR (95% CI) | p-value | aOR (95% CI) | p-value |
| **Study Arm** |  |  |  |  |
| Inpatient (vs community) | 1.79 (1.00-3.26) | 0.054 | 1.68 (0.81-3.53) | 0.164 |
| Sepsis (vs community) | **2.45 (1.48-4.12)** | **<0.001** | 1.08 (0.54-2.22) | 0.822 |
| **Demographics** |  |  |  |  |
| Age (per year) | 1.00 (0.99-1.02) | 0.709 | 1.00 (0.98-1.02) | 0.922 |
| Male sex (vs female) | 1.23 (0.84-1.82) | 0.287 | 1.44 (0.94-2.21) | 0.098 |
| **HIV status** |  |  |  |  |
| HIV+ (vs HIV-) | **1.68 (1.09-2.59)** | **0.018** | 1.21 (0.48-2.99) | 0.679 |
| HIV unknown (vs HIV-) | 0.71 (0.40-1.24) | 0.229 | 1.08 (0.54-2.16) | 0.820 |
| ART (vs none) | **1.99 (1.32-3.00)** | **0.001** | 1.07 (0.35-3.23) | 0.905 |
| CPT (vs none) | **2.46 (1.58-3.86)** | **<0.001** | 2.34 (1.00-5.66) | 0.053 |
| **Healthcare exposure** |  |  |  |  |
| Current TB treatment | 1.02 (0.33-2.99) | 0.971 | 0.51 (0.13-1.80) | 0.300 |
| Antibiotics*† | **1.81 (1.05-3.16)** | **0.034** | 1.24 (0.64-2.41) | 0.528 |
| Hospitalisation† | **7.87 (2.57-34.22)** | **0.001** | **6.64 (1.98-30.75)** | **0.005** |
| **Household** |  |  |  |  |
| Unprotected water source | 2.43 (0.96-6.64) | 0.068 | **2.96 (1.07-8.75)** | **0.040** |
| Treat water (vs not) | 1.16 (0.50-2.66) | 0.725 | 0.95 (0.37-2.37) | 0.913 |
| Flushing toilet (vs. not) | 0.72 (0.29-1.80) | 0.481 | 1.11 (0.41-3.04) | 0.842 |
| Adults (per 1) | 1.14 (0.99-1.31) | 0.064 | **1.20 (1.03-1.40)** | **0.024** |
| Children (per 1) | 1.00 (0.87-1.14) | 0.979 | 0.98 (0.84-1.13) | 0.747 |
| Keep animals (vs. not) | 1.33 (0.88-2.03) | 0.176 | 1.15 (0.72-1.84) | 0.552 |
| **Season** |  |  |  |  |
| Rainy season‡ (vs. dry) | **2.05 (1.38-3.06)** | **<0.001** | **2.21 (1.40-3.50)** | **<0.001** |

CPT = Co-trimoxazole preventative therapy, ART = antiretroviral therapy, TB = tuberculosis. Entries in bold are those for which 95% confidence intervals do not cross 1.
\* Antibiotics includes TB therapy but excludes CPT. † Hospitalisation and antimicrobial exposure within 28 days prior to enrolment. ‡ Rainy season defined as November-April

Following enrolment, there was rapid increase in ESBL-E colonisation prevalence in the antimicrobial exposed inpatient group (109/222 [49%] at day 0 to 127/162 [78%] at day 7) compared to the inpatient antimicrobial-unexposed group (41/99 [41%] at day 0 to 32/62 [51%] at day 7, Figure 1A). The most commonly received antimicrobial was, as expected, ceftriaxone (183/225, 80%) but co-trimoxazole (110/225, 49%), ciprofloxacin (61/225 27%) and antitubercular chemotherapy (52/225, 23%) were also commonly administered; because of prolonged continued CPT administration, exposure to co-trimoxazole in terms of person-days of exposure was significantly higher than for other antimicrobials (Supplementary Figure 4 and Supplementary Table 2).

We used continuous-time multi-state Markov models to understand determinants of ESBL-E carriage. In this model, each patient was assumed to exist at any one time in either a “colonised” or “non-colonised” state, with the transition rate between states governed by a linear function of time-varying covariates (hospitalisation and antimicrobial exposure). When comparing a stepwise-constant covariate model (where the effect of hospitalization and antimicrobial exposure cease immediately as exposure ceases) to a model which included a prolonged effect of antimicrobial exposure, modelled as an exponential decay that continues to exert an effect when exposure ceases, the latter was a better fit to the data as assessed by leave-one-out cross validation (estimated expected pointwise log predictive value [ELPD] difference 10.5 [standard error 4.2] in favour of the exponential decay model) and posterior predictive checks (Supplementary Figure 5). In this final model the estimated mean time in the ESBL-E colonised and uncolonised states in the absence of antimicrobial exposure or hospitalization was 9.7 (95% CrI 4.2-25.1) and 5.8 (95% CrI 2.5-14.3) days respectively. Hospitalisation significantly increased both ESBL-E gain and loss parameters resulting in a modest increase in overall carriage prevalence, whereas antibacterial therapy largely acted to prolong ESBL-E carriage by reducing loss and acted with a prolonged effect with half-life 43.7 (95% CrI 15.4-97.7) days (Figure 1C-E, Supplementary Table 3). Posterior plots of pairs of parameters revealed some non-identifiability between the gain and loss parameters, manifesting as correlation (Supplementary Figure 6).

Posterior predictive simulations from the final fitted model (Figure 1B) considering a hypothetical seven-day hospital admission with seven, two, or zero days of antimicrobial therapy suggest that antimicrobial therapy and hospitalization act in synergy to produce the observed rapid increase in ESBL-E, but that there is very little difference in ESBL-E prevalence carriage from truncating seven days of antimicrobial therapy to two days.

### Whole genome sequencing does not support horizontal gene transfer as the primary mechanism of within-participant ESBL persistence

Next we used short-read whole-genome sequencing as a high-resolution typing tool to track bacteria and ESBL genes within study participants. Following quality control, 473 *E. coli* and 203 *K. pneumoniae* species complex genomes, respectively, were included in the analysis with a median 3 (2-4) *E. coli* isolates per participant from 230 participants and 2 (1-2) *K. pneumoniae* complex isolates from 142 participants, each from a different collection time point. An analysis of population structure, core gene phylogeny and AMR gene content of these isolates has been described elsewhere^18,19^; (the majority (n= 190) of *K. pneumoniae* complex isolates were *K. pneumoniae* subsp. *pneumoniae*. To track bacteria within participant we used assemblies constructed by mapping reads to reference genomes to define sequence clusters (using popPUNK), and SNP-clusters (defined by whole-genome SNP distance of 5 or fewer SNPs). PopPUNK grouped *E. coli* into 87 clusters representing 58 sequence types (STs), and *K. pneumoniae* complex into 91 clusters representing 75 STs, 55 of these *K. pneumoniae sensu stricto*. These clusters (henceforth referred to as popPUNK clusters) were concordant with the core-gene phylogenies (Supplementary Figure 7). To track ESBL genes and their genomic environment we clustered *de novo* assembled contigs containing 3GC resistance genes using the cd-hit algorithm, including those from both *Klebsiella* and *E. coli*. 714 3GC-resistance gene containing contigs were identified in 672/676 samples; 18 different genes were carried on contigs of median length 11.9kb (IQR 4.7-102.2kb) and formed 195 clusters (henceforth, contig-clusters), of median size 1 (range 1-42, Supplementary Figure 8). They were genus and lineage associated (Supplementary Figure 9), though 21/195 (11%) of contig-clusters contained both *E. coli* and *K. pneumoniae* complex genomes.

For participants colonised with *E. coli* or *K. pneumoniae* complex at a time t=0, the probability of remaining colonised returned to a baseline by 100-150 days (Figure 2A-B), but the probability of remaining colonised with the same contig-cluster or popPUNK cluster was lower, and the probability of remaining colonised with an organism differing by five or fewer SNPs was lower still (Figure 2C-D). Sensitivity analysis varying the definition of SNP cluster from 0 to 20 SNPs did not alter these conclusions (Supplementary Figure 10). Comparing within-patient sample pairs to between-patient sample pairs the popPUNK-cluster contig-cluster combination was conserved more than either popPUNK-cluster or contig-cluster alone (Figure 2D-E), consistent with the hypothesis that within-participant persistence of ESBL, where it occurs, is caused by persistence of ESBL-containing bacteria rather than horizontal gene transfer of ESBL gene.

**Figure 2:**
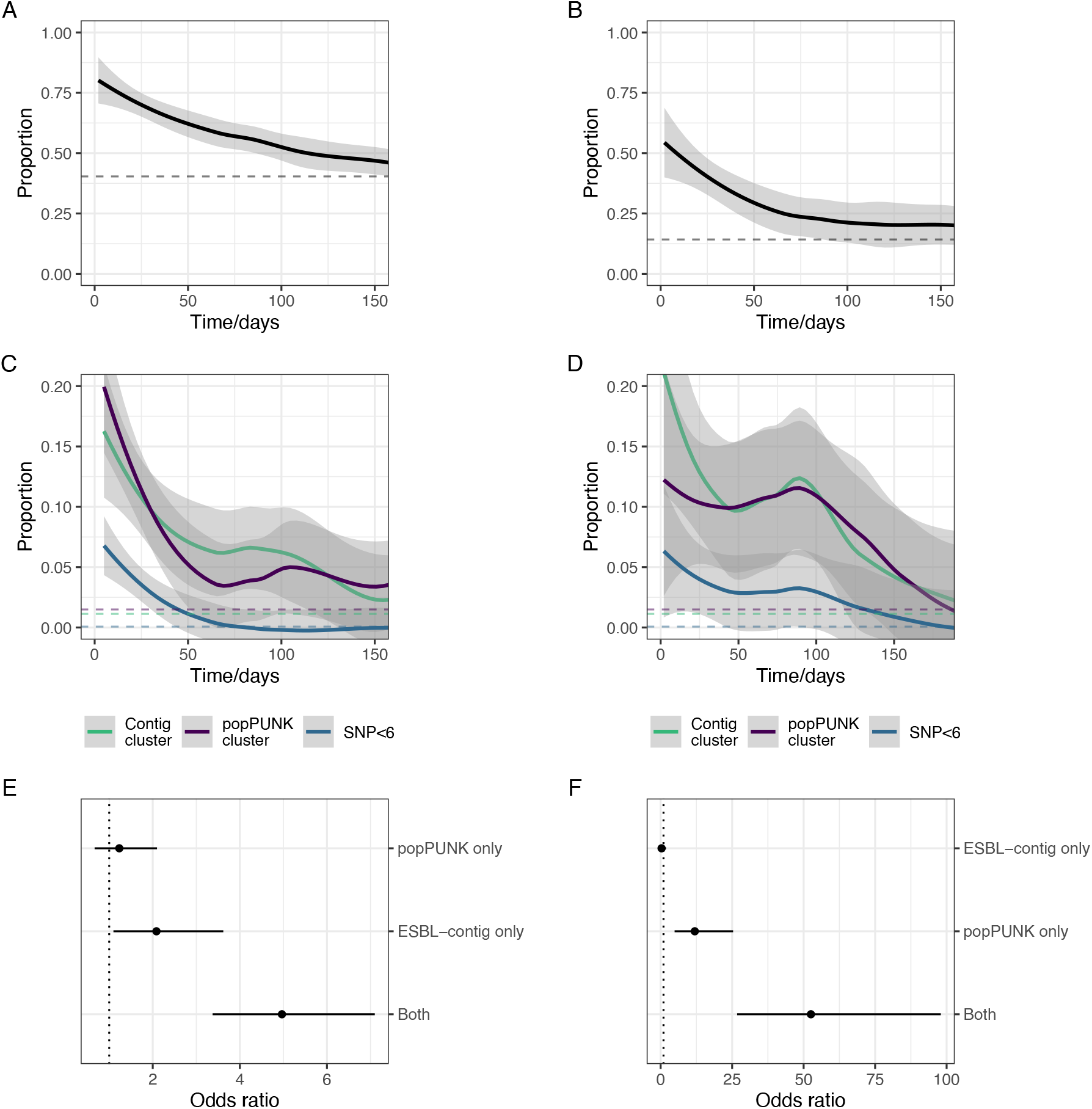
Within-participant dynamics of ESBL-E colonisation. A-B: Proportion of participants who, at time 0 have detectable ESBL producing *E. coli* (A) or *K. pneumoniae* complex (B) who remain colonised as a function of time. Dotted lines show the baseline proportion for the dataset of between-participant samples that contain the same genus. C-D: Proportion of participants with detectable ESBL producing *E. coli* (A) or *K. pneumoniae* complex (B) at time 0, who remain colonised with the same contig-cluster, popPUNK cluster, or an isolate of SNP distance ≤5 as a function of time with dotted lines showing the baseline (between-participant) proportion, as above. E-F: Odds ratio of within-participant sample pairs containing the same popPUNK cluster alone, contig-cluster alone, or both, compared to between-participant pairs, for *E. coli* (E) and *K. pnemoniae* complex (F) showing that the element that is most likely to be conserved is the popPUNK cluster-contig cluster combination.

**Figure 3:**
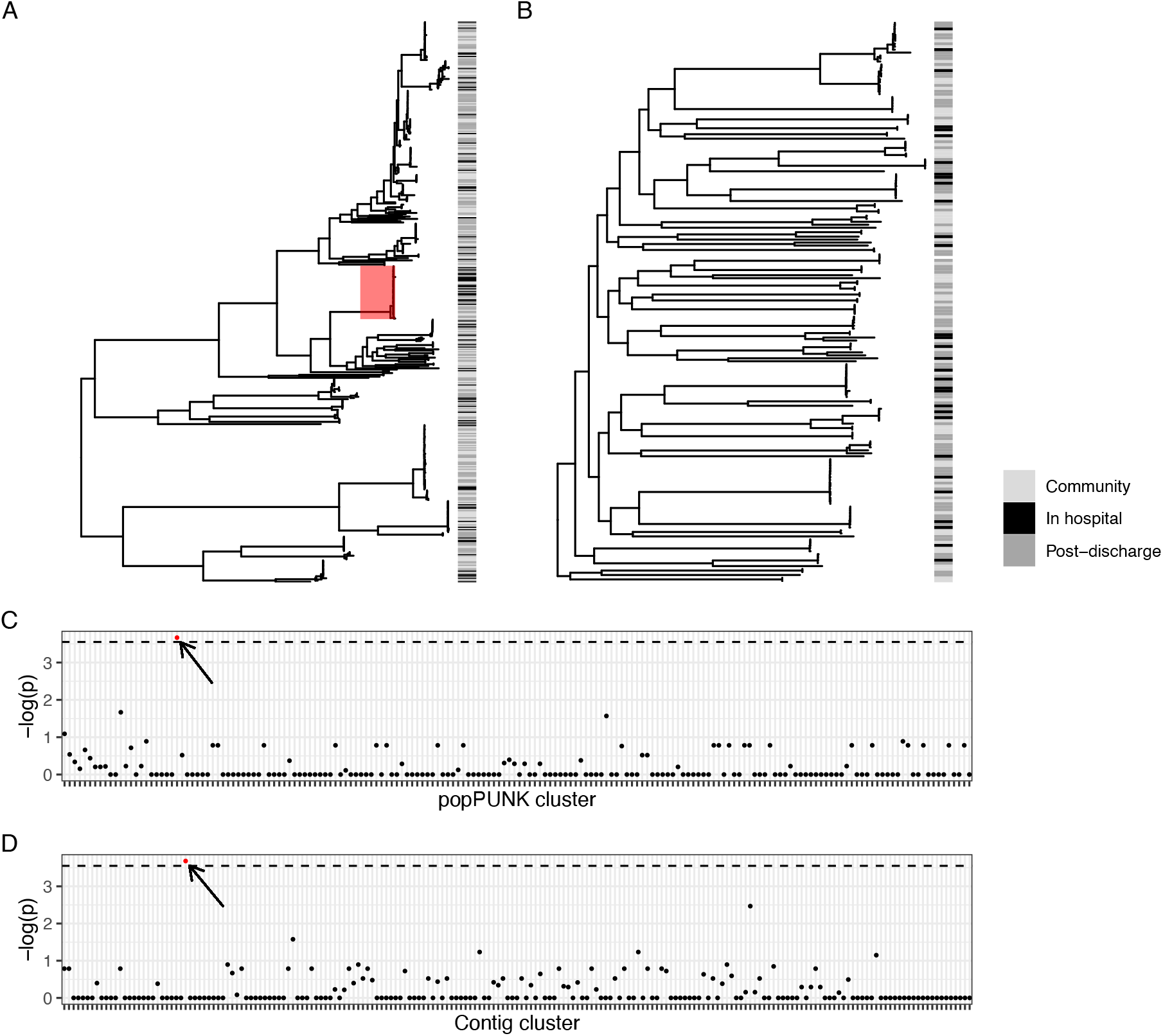
Hospital association of popPUNK clusters and contig-clusters. A-B: Maximum-likelihood core gene phylogenetic tree for *E. coli* (A) and *K. pneumoniae* subsp. *pneumoniae* (B) showing in-hospital (black), post discharge (dark grey) and community (light grey) isolates, where post-discharge is defined as up to 120 days post hospital discharge. Hospital-associated samples are distributed across the tree, but only the popPUNK cluster highlighted in red shows an association with in-hospital isolation. C-D: Manhattan plots showing p-value of Fisher’s exact test for association of popPUNK cluster (C) and contig-cluster (D) with in-hospital isolation. Dotted line shows Bonferroni-corrected value corresponding to p = 0.05. Only one popPUNK cluster is significantly associated with in-hospital isolation (highlighted in red on the plot, C, and core gene tree, A) at this level. Similarly one contig-cluster is associated with in-hospital isolation, highlighted in red; this is the contig-cluster which is associated with the hospital-associated lineage.

### Hospital associated lineages or transmission clusters are unusual

Next, we examined any hospital association of popPUNK clusters. In-hospital and post-discharge isolates were distributed throughout the core-gene phylogenies and only one popPUNK cluster contained more hospital isolates than would be expected by chance following correction for multiple comparisons (Figure 4A-C). This corresponded to *E. coli* ST410; however, 20% (9/44) of isolates belonging to this popPUNK cluster classified as community-associated highlighting that it is not exclusively hospital-associated. Similarly, one contig-cluster was associated with in-hospital isolation (Figure 4D); the *bla*_CTX-M-15_ containing contig-cluster was primarily associated with the hospital associated popPUNK cluster (Supplementary Figure 9B).

As hospital associated popPUNK clusters were unusual, we investigated putative hospital-related transmission clusters which differed by five or fewer whole genome SNPs (a “SNP-cluster”). 151/473 (32%) *E. coli* and 21/203 (10%) *K. pneumoniae* were members of a SNP-cluster and hence represent possible transmission events (Figure 5). The clusters were generally small (median size 2 [IQR 2-5] for *E. coli* and 2 [IQR 2-3] for *K. pneumoniae* complex) and, in *E. coli*, largely contained samples from different participants rather than the same participant: only 6% (10/175) of pairwise comparisons of within-SNP-cluster *E. coli* samples were from the same participant. Fewer *K. pneumoniae* formed a SNP-cluster but more were from the same participant (58% [7/12]) rather than between participants. SNP-clusters in both *E. coli* and *K. pneumoniae* complex contained community and hospital associated samples: 54/151 (35%) of *E. coli* samples that were members of a SNP-cluster were community associated, and hence had no apparent epidemiologic link, similar to the 117/321 (36%) of *E. coli* isolates that were not members of a SNP-cluster (p = 0.92 comparing prevalence of community-associated isolates using Fisher’s exact test). 4/21 (20%) *K. pneumoniae* isolates that were members of a SNP cluster were community associated, compared to 70/181 (39%) of those that were not members of a SNP cluster (p = 0.1). The choice of 5 SNPs to define a SNP-cluster is a common convention but is arbitrary; but in sensitivity analysis varying the SNP threshold from 0 to 10 did not significantly alter the conclusions (Supplementary Figures 11-13).

**Figure 5:**
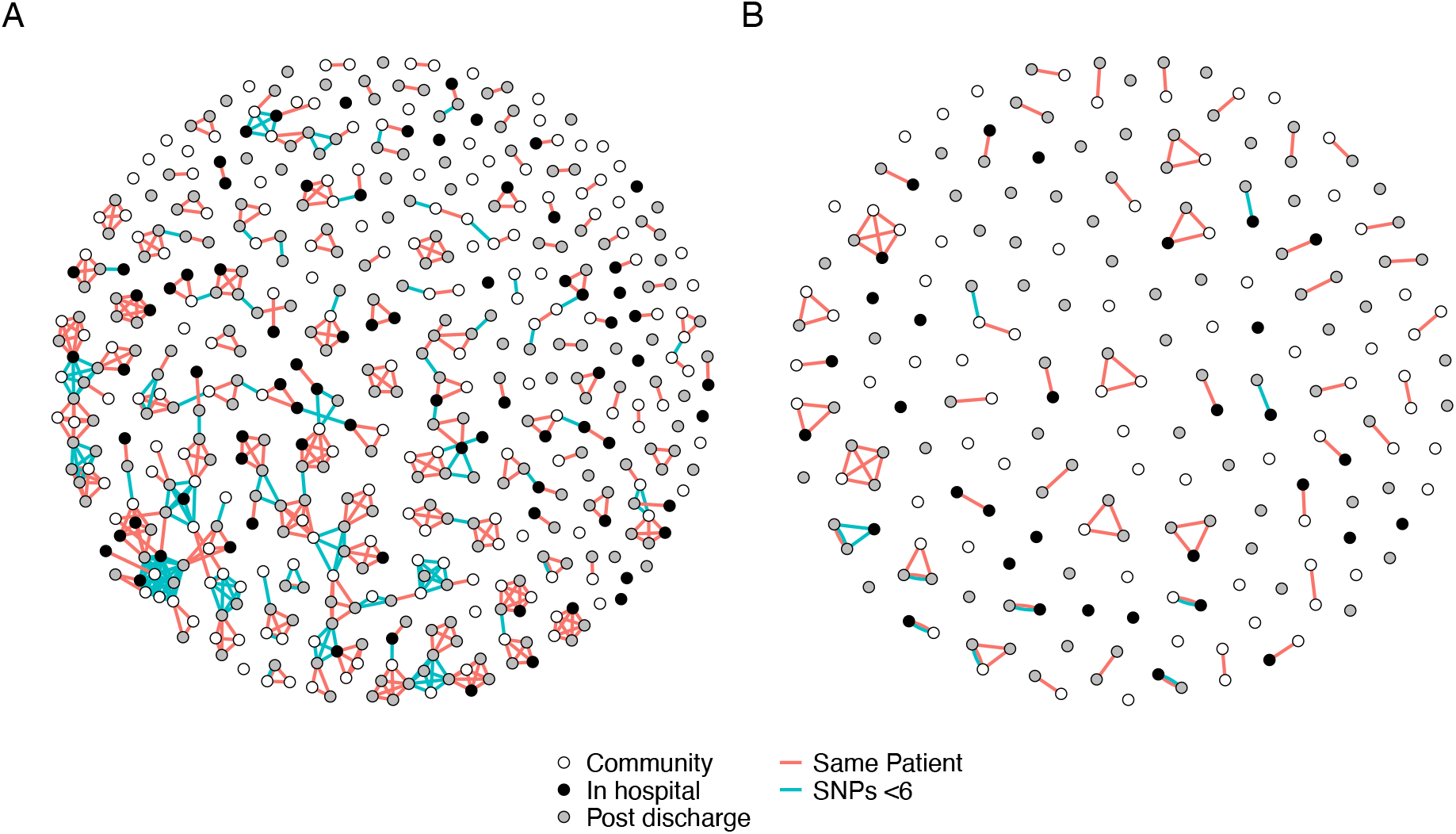
Network plot of SNP-clusters (putative transmission clusters) for *E. coli* (A) and *K. pneumoniae* (B) showing that putative transmission clusters are not exclusively hospital associated. Points are samples, coloured by place of isolation (in-hospital [black], community [white], or up to 120 days post-discharge [grey]). Red lines link samples that are within a single participant. Blue lines link samples that are differ by five or fewer SNPs. The plot shows that most samples are not members of a SNP-cluster; that most SNP clusters encompass samples from different (rather than multiple samples from the same) participants; and that SNP-clusters are not exclusively hospital-associated, i.e. they contain in-hospital, community, and post-discharge samples.

## Discussion

Combining longitudinal sampling, multi-state modelling and whole genome sequencing, we describe the dynamics of ESBL-E colonisation in Malawian adults. These findings advance our understanding of the effects of antimicrobial exposure on AMR-acquisition, with potentially significant implications for both the design of antimicrobial stewardship protocols and directions for future research.

First, baseline sampling provides insight into drivers of ESBL-E colonisation in Blantyre. ESBL-E colonisation is very common, consistent with other studies across sSA^13^. This, along with identification of community risk factors for baseline colonisation, suggest significant community spread. Observed associations of unprotected water use for drinking and higher prevalence in rainy season suggest inadequate access to water, sanitation and hygiene (WASH) infrastructure and/or WASH behavioural practices may be strongly contributing, and associations of colonisation with household crowding suggests within-household transmission may be significant.

Second, novel Markov models fitted to longitudinal sampling data allow insight into the dynamics of ESBL-E colonisation and de-colonisation. We demonstrate a rapid increase in ESBL-E colonisation following hospital admission and antimicrobial exposure. Modelling suggests that antimicrobial exposure may act to drive this increase and promote sustained carriage by exerting an effect long after antimicrobial exposure finishes, with a half-life of 43.7 (95% CrI 15.4-97.7) days.

Simulations using the fitted models suggest that, due to the sustained effect of antimicrobials, short courses of antimicrobials could exert a similar effect to prolonged courses in terms of ESBL-E carriage. This finding has clear implications for antimicrobial stewardship protocols, suggesting that truncating courses of antimicrobials may have limited effect on ESBL-E carriage compared to avoiding antimicrobial administration altogether. Previous ESBL-E longitudinal sampling and modelling studies examining the effect of antimicrobials on colonization have examined community and post-travel carriage in adults in the Netherlands^5,6,8^ and transmission of ESBL-E in neonatal units in the high prevalence setting of Cambodia^20^. In the former studies, some association of ESBL-E carriage with antimicrobial exposure was found, but antimicrobial exposure was not common, further, sampling was neither intensive nor linked to antimicrobial exposure to fully define the effects. In a Cambodian neonatal unit, antimicrobial therapy was robustly linked to an increased daily probability of acquiring *K. pneumoniae* colonisation but long-term sampling was not available to define long term antimicrobial effects as we have done here. Understanding the dynamics of ESBL-E colonisation and decolonisation under antimicrobial pressure to guide stewardship efforts should be a priority in other settings.

More broadly, these findings highlight a need to define and measure clinically relevant individual-level AMR endpoints in trials of antimicrobial treatment strategies. An expanding evidence base has demonstrated equivalence of clinical outcomes in a variety of clinical infection syndromes for shorter versus longer courses of antimicrobial therapy^21–23^, but quantifying the effect of antimicrobials on promoting AMR at the individual level is not the norm. A non-linear relationship between antimicrobial exposure and colonisation with AMR-bacteria, as we demonstrate here, may mean that seven compared to fourteen days of antimicrobials has little benefit in reducing colonisation. Defining clinically relevant AMR endpoints for trials, measuring them, and understanding their relationship with antimicrobial exposure is crucial for optimising the way in which antimicrobials are used to minimise pressure for AMR development at the individual level.

Third, using whole-genome sequencing as a high-resolution typing tool allowed us to approach the mechanism of the effect of antimicrobials in promoting ESBL-E carriage. A key question is whether the apparent rapid ESBL-E acquisition following the combination of hospital and antimicrobial exposure represents true transmission from healthcare settings facilitated by antimicrobial-induced loss of colonisation resistance, or selection for low abundance resistant bacteria that were already present in the microbiota but not detected by bacterial culture. We found limited support for hospital associated lineages or hospital-associated transmission clusters suggesting either that ESBL-E acquisition had occurred in the community and was selected for by antimicrobial exposure in hospital, or that the diversity of isolates transmitted in the hospital was contained in the diversity of isolates in the community, which is a distinct possibility in our setting. Genomic epidemiology studies of ESBL-E colonisation and infection clearly demonstrate that true healthcare associated transmission of ESBL-E occurs^15,24,25^ but few studies have longitudinal sampling pre-, during- and post- antimicrobial exposure. Defining the contribution of antimicrobial selection versus novel acquisition events in driving ESBL-E prevalence following antimicrobial exposure across high and low-income settings will guide prevention efforts and should be a priority for future studies. Healthcare associated transmission could be reduced by infection prevention and control procedures, but antimicrobial selection pressure would need novel strategies to protect the microbiota against selection for resistant bacteria such as antimicrobial binding compounds^26^ or oral beta lactamases^27^.

We demonstrate considerable within-participant ESBL-E bacterial diversity (as defined by SNP- and popPUNK-clusters) over time even in participants who remain colonised with the same genus; a further key question is whether this temporal bacterial diversity with preserved ESBL-E colonisation could represent horizontal gene transfer of ESBL genes between bacteria. Horizontal gene transfer could also explain an apparent lack of hospital-associated transmission clusters, if ESBL genes disseminated into diverse clones in the healthcare setting. We find that within-participant the popPUNK-cluster contig-cluster combination was conserved more than either popPUNK-cluster or contig-cluster alone, consistent with the hypothesis that within-participant persistence of ESBL, where it occurs, is caused by persistence of ESBL-containing bacteria rather than horizontal gene transfer of ESBL genes to differing bacterial hosts. This does not support the hypothesis of horizontal gene transfer as primary mechanism of ESBL temporal persistence within-participant on the timescale of the study.

There are limitations to our study. Most importantly, due to resource limitation, we took only one colony pick from each patient-time point sample for sequencing and so we may have missed intra-host ESBL-E diversity. Within-participant ESBL-E diversity was considerable over time, and we are unable to say whether this represents frequent colonisation and de-colonisation or sampling of within-host diversity; previous studies have shown that some ESBL-E colonised participants harbour significant ESBL-E diversity^28^, so the latter is probable. Even so, if the single colony pick represented an unbiased sampling of one ESBL-E at each time point, our observations regarding temporal trends should remain valid. We used short read sequencing and clustered ESBL-containing contigs as a proxy for mobile genetic elements (MGEs), which is likely to have introduced some error: short read sequencing and de-novo assembly is usually unable to fully assemble MGEs such as plasmids, upon which many ESBL genes would be expected to be carried. It is possible that short partially assembled ESBL-contigs representing (for example) common ESBL-E containing transposons could be matched to longer plasmid fragments where the same transposon is carried on a different plasmid backbone.

Also some ESBL genes are likely to be chromosomally integrated. Long read sequencing could provide the resolution to fully describe MGEs but was not carried out due to resource limitation. We used an arbitrary SNP threshold of 5 SNPs to define SNP-clusters, a strong assumption; this cut-off (empirically derived) has been used by public health bodies in England and Canada to define possible *E. coli* outbreaks^29,30^; other authors have suggested a cut-off of 10 or below^31^. Similar empirically derived SNP cut-offs of 7-12 have been suggested for K. *pneumoniae* complex^32–34^. Nevertheless this assumption could misclassify isolates, a risk we have tried to mitigate with sensitivity analysis. We did not account for temporal distance between samples though variation in SNP distance due to acquisition of mutations over the course of the study (18 months) would be expected to be small based on experimentally determined rates of mutation acquisition^35^. We used a map-to-reference approach to identify core-genome SNPs that could have introduced bias due to the choice of reference. We have looked at high-level clustering with popPUNK and it may be that a high-resolution clustering approach using local, lineage-specific references would give the resolution to identify more hospital associated transmission events. The models of AMR carriage assumed a 100% sensitivity and specificity of sampling, which may not be valid. We were not able to disaggregate the effect of different antimicrobial agents because of the sample size and so all were treated equivalently in the models, but it is likely that there is a differential effect on ESBL-E colonization between antimicrobial classes and agents. The data do not allow us to comment on the generalisability of our modelling findings to other settings, including high income countries.

In conclusion, we describe the dynamics of ESBL-E colonisation in Malawian adults as they are exposed to antimicrobial therapy and hospitalization. Antimicrobial therapy acts rapidly to promote ESBL-E colonisation via a prolonged effect which means that truncated courses of antimicrobials may have a similar effect to longer ones, which has implications for stewardship protocols. Short-read whole-genome sequencing did not identify widespread hospital associated lineages or hospital-associated transmission clusters suggesting either that ESBL-E acquisition had occurred in the community and was selected for by antimicrobial exposure in hospital, or that the diversity of isolates transmitted in the hospital was contained in the diversity of isolates in the community. Future work should define dynamics of intra-host ESBL-E diversity under antimicrobial pressure, using longitudinal sampling, metagenomic sequencing methods to describe diversity and long-read sequencing to characterize MGEs. This will facilitate development of clinically relevant AMR endpoints for clinical trials and the development of a sound evidence base for stewardship protocols at the individual level – an evidence base which is currently lacking.

## Methods

### Study setting and design

The study took place in Queen Elizabeth Central Hospital (QECH), Blantyre, Malawi, a government tertiary referral hospital for the Southern Region of Malawi which provides free healthcare to the ∼800,000 residents^36^ of urban Blantyre. Adults (> 15 years) with sepsis, defined by fever and organ dysfunction criteria, were recruited from the emergency department of QECH 0700-1700 Monday to Friday as part of a study of sepsis aetiology, as described elsewhere^37^. Two comparator cohorts of participants were recruited: age and sex matched adults from QECH emergency department who had a plan from their attending clinical team to admit to hospital but no plan for antimicrobial administration; and community members matched by age, sex and home location to recruited sepsis patients. Exclusion criteria for the latter two groups were antimicrobial exposure within the past four weeks (except co-trimoxazole preventative therapy [CPT] and antituberculous chemotherapy); hospitalised participants who lacked capacity to give informed consent and had no guardian to give proxy consent; participants who spoke neither English nor Chichewa; and participants who lived > 30km from Blantyre city. Geographic matching on home location between community members and sepsis patients was achieved by random walk from the houses of sepsis participants with initial direction established by spinning a bottle on the floor. Written informed consent was obtained from all participants. An admission questionnaire was administered to all participants at enrolment and hospitalised patients were reviewed daily by a study team member until discharge to extract details of antimicrobial therapy from the clinical record. All clinical decisions were at the discretion of the attending clinical team. Further review by the study team occurred at day 7, 28, 90 and 180, except for community members in whom the day 7 and 90 visits were omitted. If participants failed to come to their scheduled visits, then they were traced by telephone or, if that failed, by home visit. Hospitalised patients were not financially compensated for their time, but all other participants were at a rate of 500MWK for home visits and 2000MWK for hospital visits. Data were captured using a combination of direct electronic data entry by study team members onto tablet devices (ODK^38^, Get ODK inc. United States) and paper forms (TeleForm, Opentext, Canada).

### Ethical and data availability statements

The study was approved by the research ethics committees of the Liverpool School of Tropical Medicine (16-062) and Malawi College of Medicine (P.11/16/2063). All data and code to replicate this analysis are available as the *blantyreESBL* R v1.0.0^39^ package at https://github.com/joelewis101/blantyreESBL.

### Microbiologic methods

At each study visit (enrolment, day 7, 28, 80 and 190 for hospitalised participants and enrolment, day 28 and 190 for community members) stool was collected in a sterile polypropylene pot; if a participant was not able to provide a stool sample, then a rectal swab was taken by a trained study team member and stored in Ames’ medium for transport. Stool and rectal swab samples were stored at 4°C before being batch processed weekly: samples were plated directly onto commercially available ESBL selective chromogenic agar (CHROMagar ESBL, CHROMagar, France) and cultured aerobically overnight. Morphologically distinct white or blue colonies were speciated with the API 20E system (Biomerieux, France); pink colonies were identified as *E. coli*. ESBL production was confirmed with the combination disc method on iso-sensitest agar with discs of cefotaxime (30 micrograms) and ceftazidime (30 micrograms) with and without clavulanic acid (10 micrograms), with ESBL production confirmed if there was a difference of 5mm or more between the clavulanic acid and non-clavulanic acid discs for either cephalosporin. For organisms likely to carry a chromosomal *bla*_*ampC*_ beta-lactamase gene and hence able to hydrolyse cefotaxime and ceftazidime (defined for our purposes as *Enterobacter* spp., *Citrobacter freundii, Morganella morganii, Providencia stuartii, Serratia* spp., *Hafnia alvei*); cefipime (30 micrograms), an AmpC-stable cephalosporin was used with and without clavulanic acid (10 micrograms), and ESBL production confirmed if there was a difference of 5mm or more between the clavulanic acid and non-clavulanic acid discs.

### DNA extraction, sequencing and bioinformatic analysis

One of each morphologically distinct *K. pneumoniae* species complex and *E. coli* colony, respectively, from each patient at each time point was taken forward for DNA extraction and whole-genome sequencing. DNA was extracted from overnight nutrient broth culture using the Qiagen DNA mini kit as per the manufacturer’s instructions. Extracted DNA was shipped to the Wellcome Sanger Institute to undergo whole-genome sequencing using Illumina HiSeq X10 to produce 150bp paired end reads. Quality control, de-novo assembly and construction of core gene phylogeny are described elsewhere^18,19^; in brief, species was confirmed with Kraken v0.10.6 and Braken v1.0^40^ before de-novo assembly with SPAdes^41^, with the modifications described by Page et al.^42^ and annotation with prokka v1.5^43^ using a genus specific database from RefSeq. The Roary v1.007 pan-genome pipeline^44^ were used to identify core genes, considering genes contained in at least 99% isolates to be core. Samples with assembly failure (< 4Mb assembled length) and samples with > 10% contamination (as defined by CheckM v1.1.3^45^) were excluded from the analysis. A core gene multiple sequence alignment was generated using mafft v7.205^46^, SNP-sites identified using SNP-sites v2.4.1^47^ and the resultant SNP alignment (99,693 variable sites from a core gene alignment of 1.39Mb bases for *E. coli* and 378,596 variable sites from a 2.82Mb core gene alignment for *K. pneumoniae* complex) used to infer a maximum-likelihood phylogenetic tree using IQ-TREE v1.6.3^48^ with ascertainment bias correction and the ModelFinder module, which selected the generalised time reversible model with FreeRate heterogeneity with 5 parameters for *E. coli* and 8 parameters for *K. pneumoniae* complex. 1000 ultrafast bootstrap replicates were generated. Trees were visualized with ggtree v2.2.4^49^.

AMR genes and plasmid replicons were identified using ARIBA^50^ and the curated ARG-ANNOT database used by SRST2^51^ and PlasmidFinder^52^ databases, respectively, on the sequence reads. ARIBA was also used to identify multilocus sequence type (ST) using the 7-gene *Klebsiella*^53^ and 7-gene Achtman^54^ *E. coli* schemes hosted at pubMLST (https://pubmlst.org/). To track putative mobile genetic elements within-participant over time we clustered ESBL-containing contigs from the *de novo* assemblies (identified with BLAST^55^ using the SRST2 database) to form contig-clusters using cd-hit-est v4.8.1^56^ with 95% sequence identity and otherwise default settings. To track bacteria within-participant we used popPUNK^57^ on mapped assemblies: we used snippy v4.6.0^58^ to map reads to K-12 MG1655 *E. coli* (ENA accession U00096) and MGH78578 *K. pneumoniae* (ENA accession GCA_000016305.1) references. The *E. coli* mapped assemblies had a mean (SD) coverage and depth of 92% (2%) and 58x (8x) respectively and the *K. pneumoniae* complex 92% (3%) and 52x (16x). We then used popPUNK v2.0.2 on these assemblies, forming a new database with minimum kmer size 15 (and otherwise default settings) and clustering with the DBSCAN algorithm. To compare SNP distances between samples, we used these snippy-generated assemblies (i.e. using reads mapped to the references above) to construct a multiple sequence alignment), filtered regions of presumed recombination with gubbins^59^ and calculated pairwise SNP distances using snp-dist v0.6.2 (https://github.com/tseemann/snp-dists) and considered two isolates with 5 or fewer SNPs difference across the genome to be likely to represent the same isolate. We hence used this SNP difference to define a “SNP-cluster”, clustering isolates with hierarchical clustering using the function *stats:*:*hclust* in R. We performed sensitivity analysis and varied this SNP threshold from 0 -10.

### Statistical analysis

All statistical analyses were carried out in R v4.0.2 (R foundation for statistical computing, Vienna, Austria). Summaries of variables are presented as proportions (with exact binomial confidence intervals where appropriate) or medians with interquartile ranges. Kruskal-Wallace and Fisher’s Exact tests were used to test the equivalence of patient characteristics across the three study groups for continuous and categorical variables, respectively. Associations of baseline ESBL-E carriage were assessed using logistic regression, including all variables that were felt *a priori* to be associated with ESBL-E carriage as predictors, and presenting results as odds ratios for predictor variables with 95% confidence intervals.

To assess within-participant conservation of organism, popPUNK cluster, contig-cluster, and SNP-cluster, we plotted within participant correlation curves, including all participants who were colonised with *E. coli* or *K. pneumoniae* at time t = 0 then using non-parametric LOESS regression as implemented in the R *stats::loess* function with parameters n= 80, span = 0.75 to estimate the proportion at a time t later who were colonised with the same organism, popPUNK cluster, contig-cluster, or SNP-cluster. To assess the probability of two within-participant samples containing the same cluster by chance we compared the within-participant cluster conservation proportion to the proportion of between-sample participants that contained the same cluster. Odds ratios with 95% confidence intervals were used to assess the odds of within-participant conservation of popPUNK cluster and contig-cluster together or each alone compared to between-participant conservation.

We assessed for hospital associated lineages by mapping metadata to the core gene trees, defining isolates as either in-hospital (if they were isolated from a sample taken in hospital) recent discharge (if they were isolated from a sample taken up to 120 days following hospital admission) or community (if they were neither in-hospital nor recent discharge). We tested the hypothesis that popPUNK and contig clusters are healthcare associated by comparing the proportion of healthcare-associated isolates (defined as in-hospital or recent discharge) for each cluster to the proportion of the remaining samples, using a Bonferroni-corrected Fisher’s exact test.

We looked for putative transmission clusters by plotting SNP clusters using the R packages *igraph*^60^ and *ggraph*. We used Fisher’s exact test to compare the proportion of isolates that were community-associated between isolates that were members of a SNP-cluster and those that were not.

## Modelling of ESBL-E carriage

### Defining the likelihood of the model

To understand the dynamics of ESBL-E carriage, we extended the continuous time Markov models as implemented in the MSM^17^ package in R. MSM allows stepwise constant time-varying continuous time Markov models, whereas we aimed to assess the biologically plausible effect of allowing antimicrobial exposure to act with a non-stepwise time-varying effect.

We assumed a two-state system with *N* participants, where at time *t* participant *n* will be in a state *S*_*n*_(*t*) – either ESBL-E colonised (*S*_*n*_(*t*) = 1) or ESBL-E uncolonised (*S*_*n*_(*t*) = 0). For each participant *n* we assume have a measured value of *S*_*n*_(*t*) at *i*_*n*_ time points, the times of which are given by 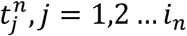, and so the *i*_*n*_ values of 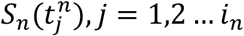 are known.

If we develop a model with parameters *θ* that predicts the probability of a particular participant being in a state *S*_*n*_(*t*_*b*_) at a time point *t*_*b*_ given that they were in a state *S*_*n*_(*t*_*a*_)at an earlier time point then then the likelihood of this observation is:

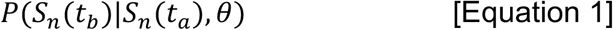

Where | indicates “conditional on” as per standard probability notation. Assuming that all observations are independent then the likelihood for any participant is the product of all the transitions for that participant; and the likelihood of the data we observe is the product of all transitions for all participant:

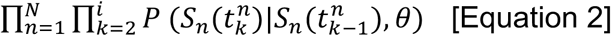

We assume a Markov model as the data-generating process, where the instantaneous probability of transition from a state *i* to state *j* is given by *q*_*ij*_, or traditionally in matrix notation as the Q-matrix^17,58^ (for a two state system):

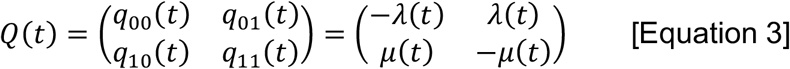

Where we have defined *λ*(*t*) as the instantaneous rate of ESBL-E loss, and *μ*(*t*) as the instantaneous rate of ESBL-E gain, and used the fact that the rows of the Q-matrix must sum to one (i.e. every participant has to be in one state or another). If we define the probability of a participant being in a state *i* at time 0 and a state *j* a time *t* as *p*_*ij*_ (*t*) = *P*(*t*), then these probabilities are linked to the Q-matrix by the set of differential equations:

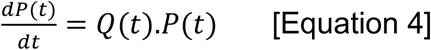

Or, simplified if participants start in a state 0 or 1 to:

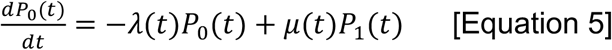

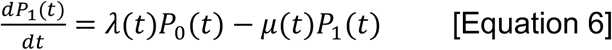

Where *P*_*i*_(*t*) is the probability of being in state *i* at time *t*. These differential equations can be solved with numerical ordinary differential equation (ODE) solvers for all state transitions and all patients to calculate the likelihood.

### Incorporating covariates

Following *msm* and Marshall and Jones^59^ we incorporated covariates with a proportional hazard approach where the *k* covariates *x*_*k*_ *k* = 1,2 …*k* can act upon the hazard of transition via:

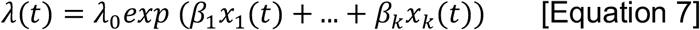

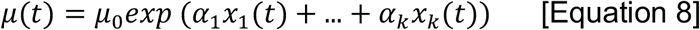

Where the *x*_*k*_ take the value 0 when an exposure is absent and 1 when it is present – this is the stepwise constant model. *λ*_0_ and *μ*_0_ are the instantaneous rate of ESBL-E loss, and the instantaneous rate of ESBL-E gain, respectively, with all covariates set to 0. The parameters *β* and *α* can therefore be thought of as the log transform of the hazard ratio of ESBL-E loss and gain, respectively; and the parameters *λ*_0_ and *μ*_0_ can be interpreted as the reciprocal of the mean time in the uncolonised or colonised state respectively with all covariates set to 0.

Finally, the motivation for developing this model was to allow a time-varying effect of antimicrobial exposure. Assuming that antimicrobial exposure begins at time *t*_*start*_ and ends at *t*_*end*_, the value of the covariate *x*_*antimicrobial*_(*t*) takes the form of an exponential decay following exposure:

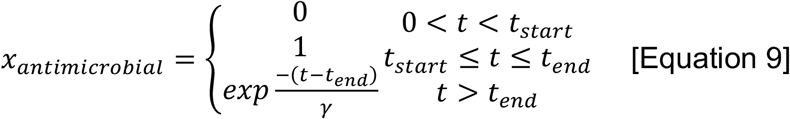

Where the parameter γ is the half life of the decay of antimicrobial exposure effect, multiplied by the natural log of 2.

### Fitting and comparing models

The models were coded and fit in a Bayesian framework in Stan v2.19^61^ accessed via the *Rstan* v2.19.2 interface in R, and plotted using the *bayesplot* v1.8 R package. All code and data to fit the models is contained in the blantyreESBL^39^ v1.0.0 R package available at https://github.com/joelewis101/blantyreESBL. Weakly informative priors were used; a normal distribution with mean 0 and standard deviation 2 for alpha and beta (corresponding (corresponding to a hazard ratio of 7.4), a normal distribution with mean 0 and standard distribution 0.2 for mu and lambda and a normal distribution with a mean of 0 and standard deviation of 50 days for gamma. In each case models were fit with four chains of 1000 iterations each with 500 warmup iterations. Convergence was evaluated by inspection of traceplots and the Gelman-Rubin statistic^62^ being close to 1. Posterior estimates of parameters were expressed as medians with 95% credible intervals generated from the quantiles of the posterior, excluding warmup iterations. We fit two models: one with the stepwise-constant covariates and one with exponentially-decaying being close to 1. Posterior estimates of parameters were expressed as medians with 95% credible intervals generated from the quantiles of the posterior, excluding warmup iterations. We fit two models: one with the stepwise-constant covariates and one with exponentially-decaying effect of antimicrobial exposure.

To compare between the two models we used leave one out cross validation as implemented in the *loo* v2.1.0 package in R^63^, quantifying model fit with an estimate of the expected log predictive density (ELPD) and comparing models with the ELPD difference and standard error of the difference, where a difference in ELPD of greater than two times the standard error of the difference could be interpreted as evidence in favour of the better fitting model^63^. We also used graphical posterior predictive checks, simulating the predicted prevalence of ESBL-E across the three arms of the study by generating a probability of ESBL-E carriage for each participant at each time point for each posterior samples (excluding warmup draws) and sampling from a Bernoulli distribution using the predicted probability. We simulated from the posterior by fixing covariate values, assuming a baseline prevalence of 50% ESBL carriage at t=0 and using all posterior draw covariate values (excluding warmup draws) and solving the likelihood differential equations using the R package *deSolve* v1.28^64^ to generate daily predicted probabilities of carriage at time t, with 95% confidence intervals defined by simple quantiles.

## Supporting information

Supplementary results

## Data Availability

All data and code to reproduce the analysis in this manuscript are available at

https://joelewis101.github.io/blantyreESBL/

## Author contributions

Conceptualisation: JL, NT, NAF, BF, CJ. Methodology: JL, NT, NAF, MAB, EH, JM, CJ. Investigation: JL, MM, RB. Formal analysis: JL, NT, NAF, EH, MAB, CJ, BF. Writing – original draft preparation; JL. Writing – review and editing: JL, MM, RB, MB, JM, EH, NT, NAF, CJ, BF. Supervision: NAF, NT

## Acknowledgements

Many thanks to the study team: Lucy Keyala, Tusekile Phiri, Grace Mwaminawa, Witness Mtambo, Gladys Namacha, Monica Matola; to the MLW laboratory teams, particularly Brigitte Denis; and to the MLW data team, particularly Lumbani Makhaza and Clemens Masesa. The authors acknowledge the sequencing team at the Wellcome Sanger Institute, and Christoph Puethe and the Pathogen Informatics team for computational support. The authors have no conflicts of interest to declare.

