## Supplementary results for "Dynamics of gut mucosal colonisation with extended spectrum beta-lactamase producing Enterobacterales in Malawi"

### Supplementary material

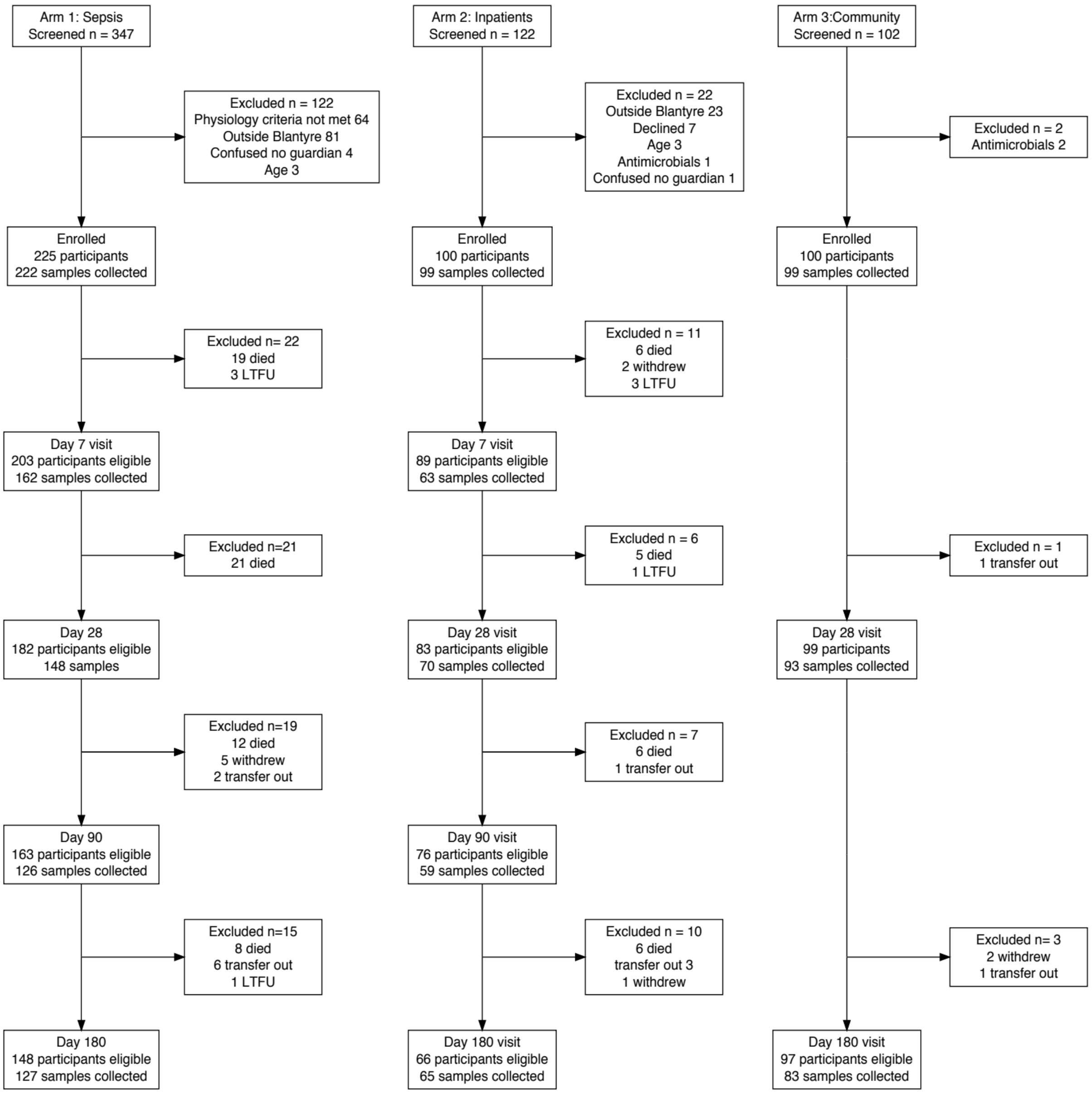


Supplementary Figure 1: Flow through the clinical study. LTFU = lost to follow up

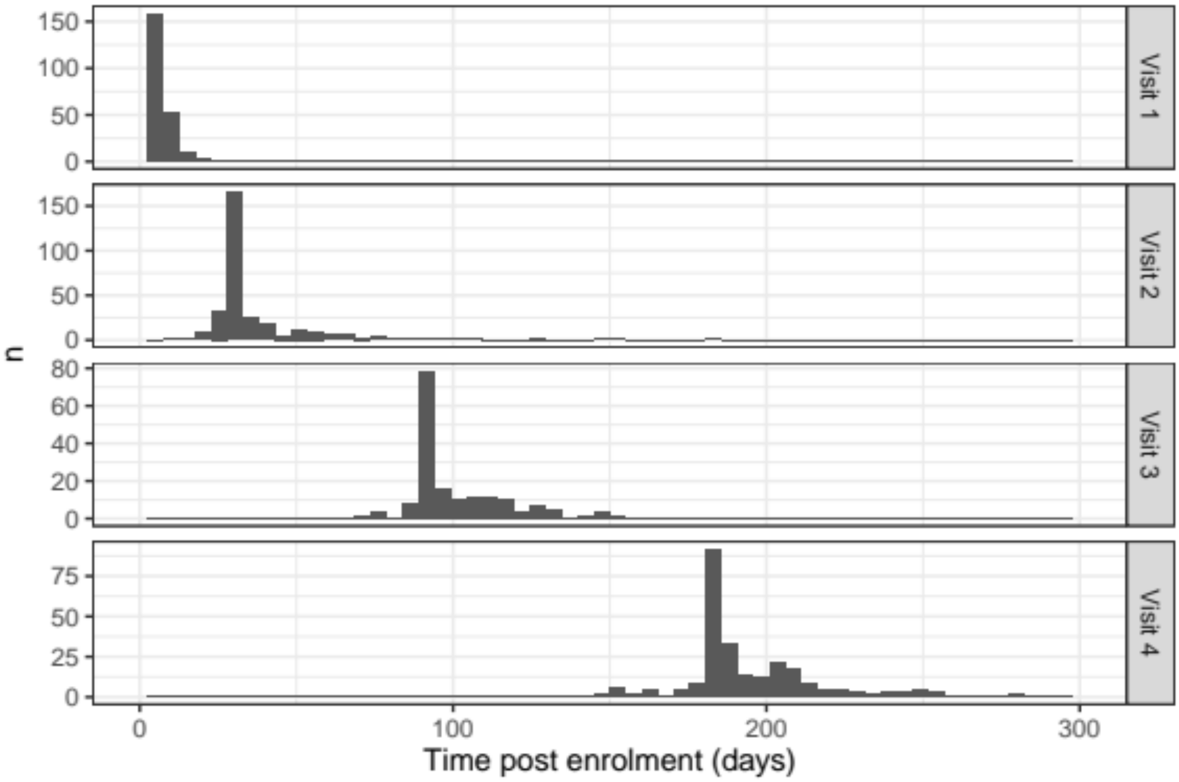


**Supplementary Figure 2:** Distribution of stool sample collection dates

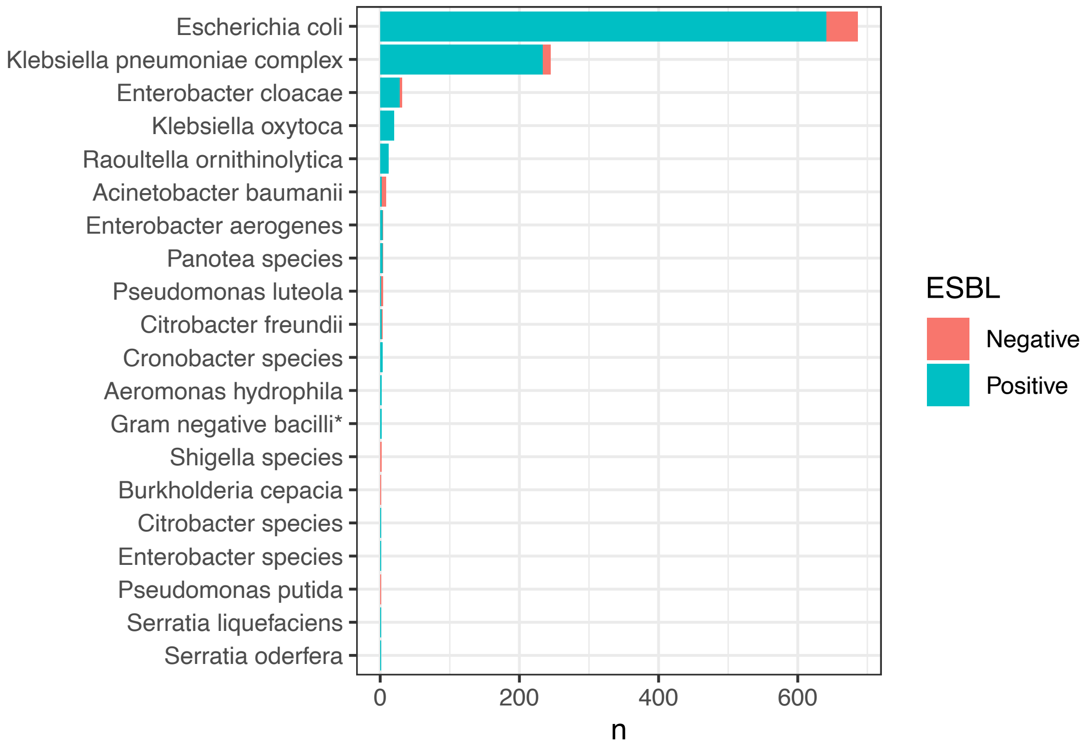


**Supplementary Figure 3:** Species of bacteria isolated from stool. Samples were labelled “Gram negative bacilli” is they could not be speciated using the API system.

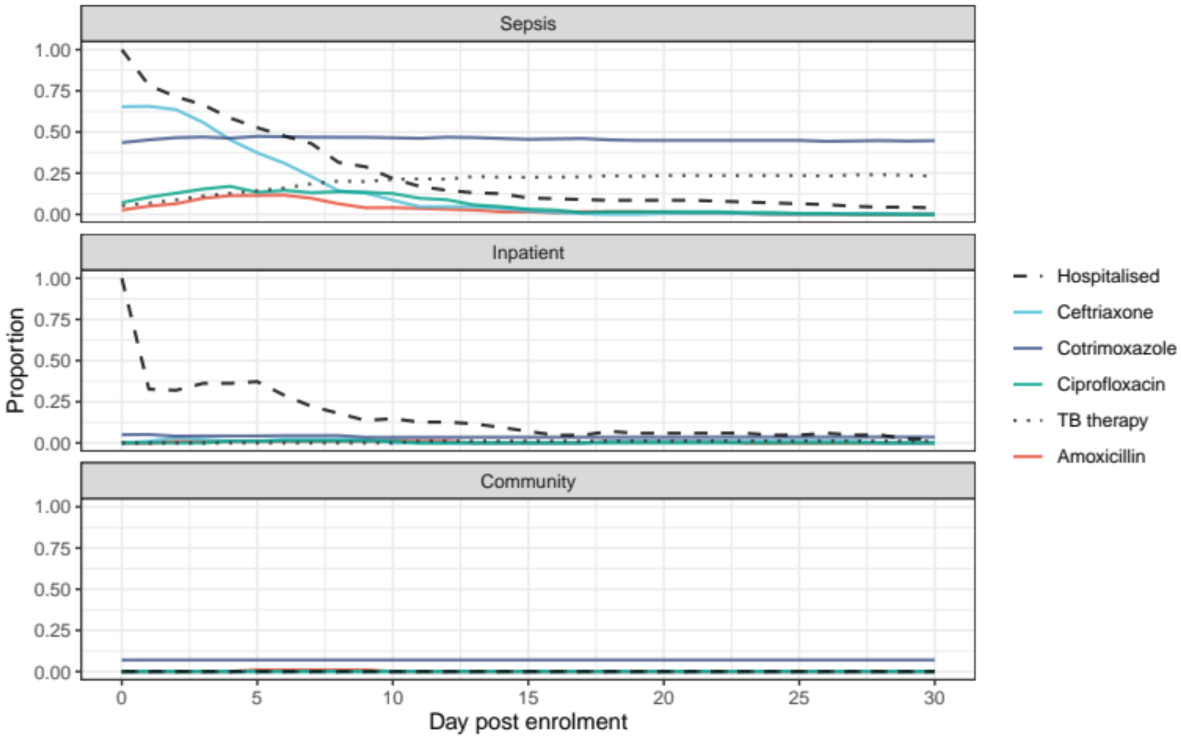


**Supplementary Figure 4:** Participant antimicrobial exposure and hospitalisation stratified by study arm.

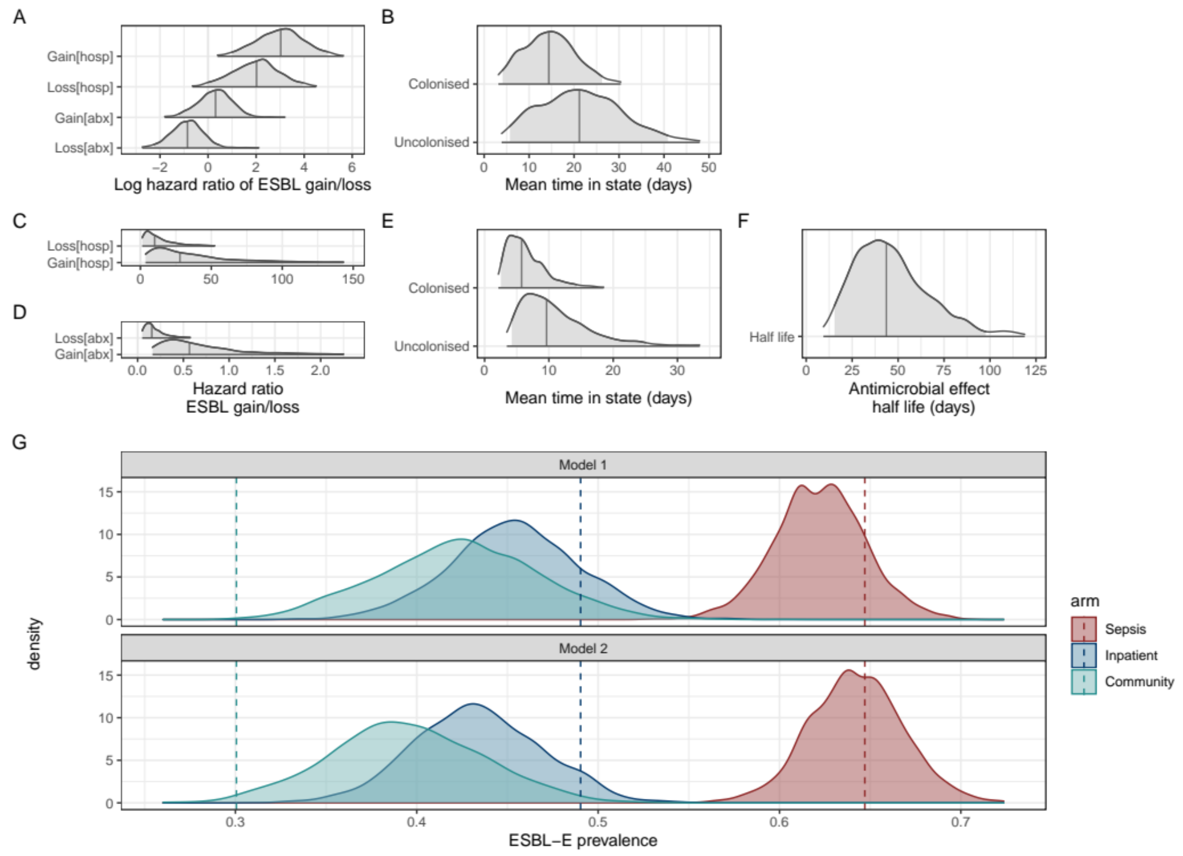


**Supplementary Figure 5:** Comparing a model of ESBL-E carriage that include a stepwise constant effect of hospitalisation and antimicrobial exposure (where the effect of covariates ceases when exposure ceases in Model 1) to a model that allows the effect of antimicrobial exposure to persist when exposure finishes, modelled as an exponential decay in Model 2. A-B: Parameter estimates from Model 1 expressed as natural logarithm of hazard ratio of gain or loss of ESBL-E for antimicrobial exposure [abx] and hospitalisation [hosp] (A) and mean time in the colonised or uncolonised state (B) with covariates set to 0 (i.e. no antimicrobials, not hospitalised). C-E: Parameter estimates from Model 2, with the same interpretation and the addition of the half-life (in days) of the decaying effect of antimicrobial exposure (E). F: Posterior parameter checks of two models showing actual prevalence of ESBL-E carriage stratified by study arm (dashed lines) with kernel density plots of predicted prevalence from fitted models, obtained by using all posterior parameter estimates (n=2000, discarding warmup iterations) to predict probability of ESBL-E from the actual data., and sampling from a binomial distribution using this probability. Model 1 underfits the antimicrobial-exposed arm of the study, which is improved by the addition of the prolonged effect of antimicrobials.

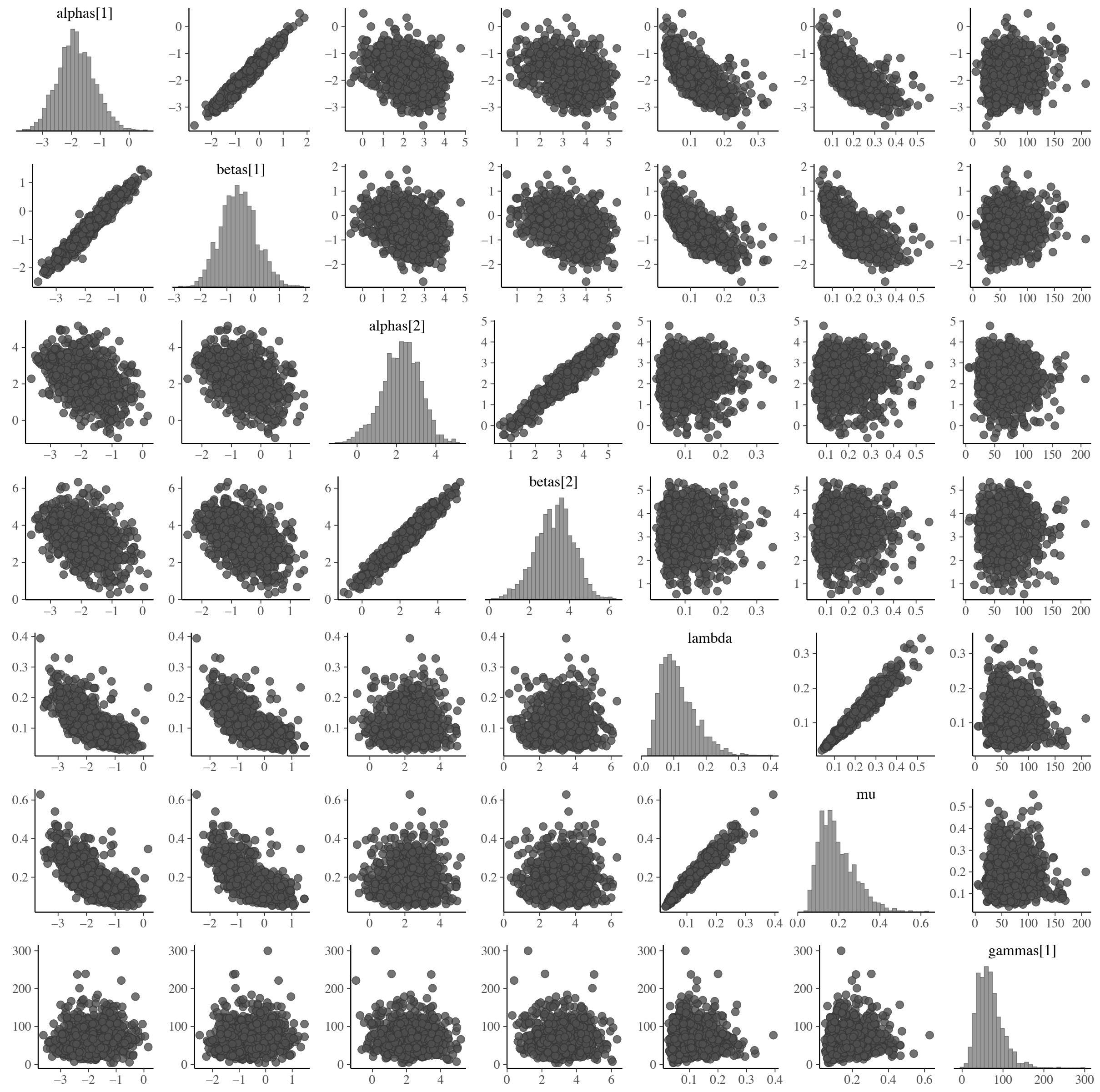


**Supplementary Figure 6:** Pairs plot of posterior parameter estimates showing nonidentifiability of gain and loss parameters manifesting as strong correlation. Alpha[1] and beta[1] are log hazard ratio of hospitalisation and alpha[2] and beta[2] of antimicrobial exposure. Lamda and mu are baseline instantaneous ESBL-E loss and gain and gamma is the scaled half-life of the effect of antimicrobial exposure.

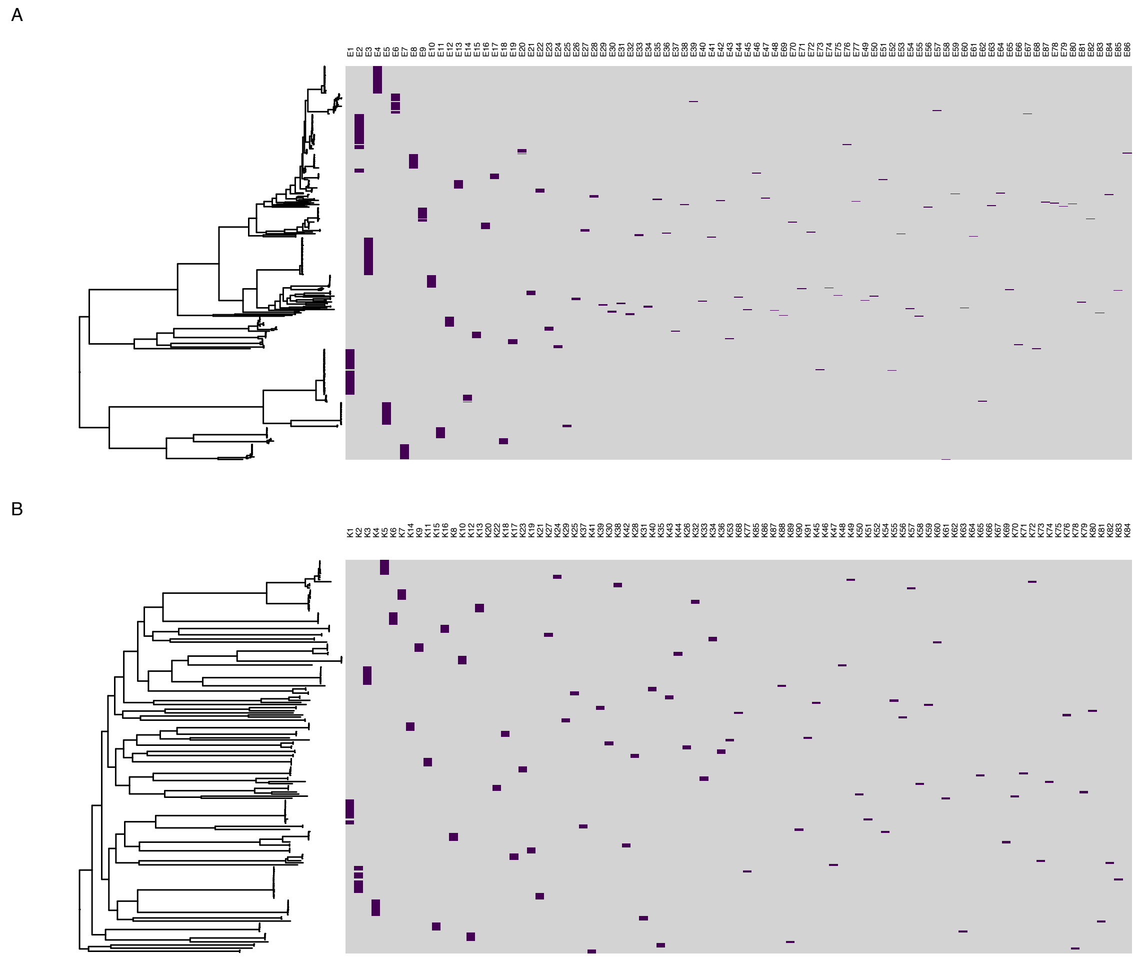


**Supplementary Figure 7:** popPUNK sequence clusters across the core-gene maximum-likelihood phylogeny for *E. coli* (A) and *K. pneumoniae* subsp. *pneumoniae* (B). popPUNK sequence cluster name shown at top of heatmap and purple indicates cluster membership. The sequence clusters correspond well to lineages inferred from the phylogeny.

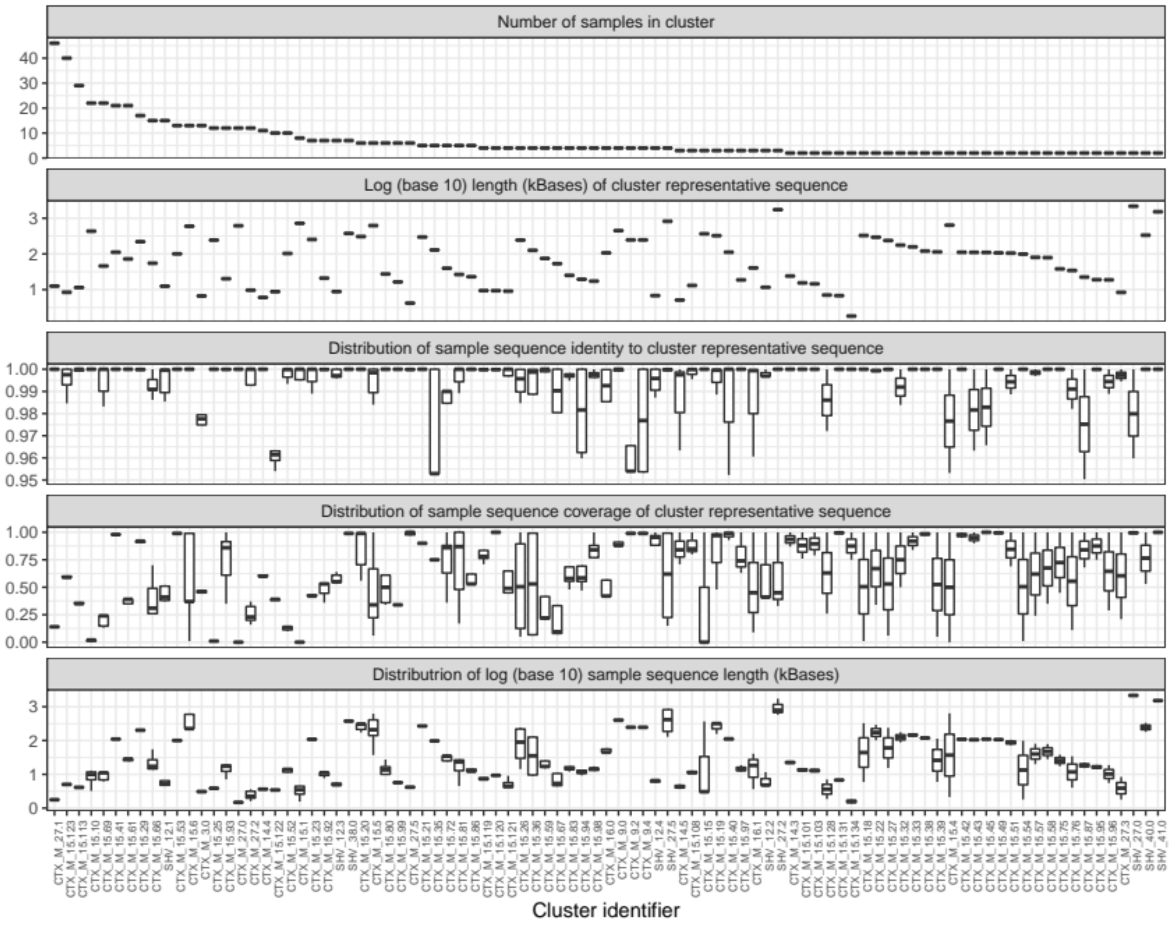


**Supplementary Figure 8:** Descriptive statistics of third-generation cephalosporin (3GC) resistance gene contig-clusters.

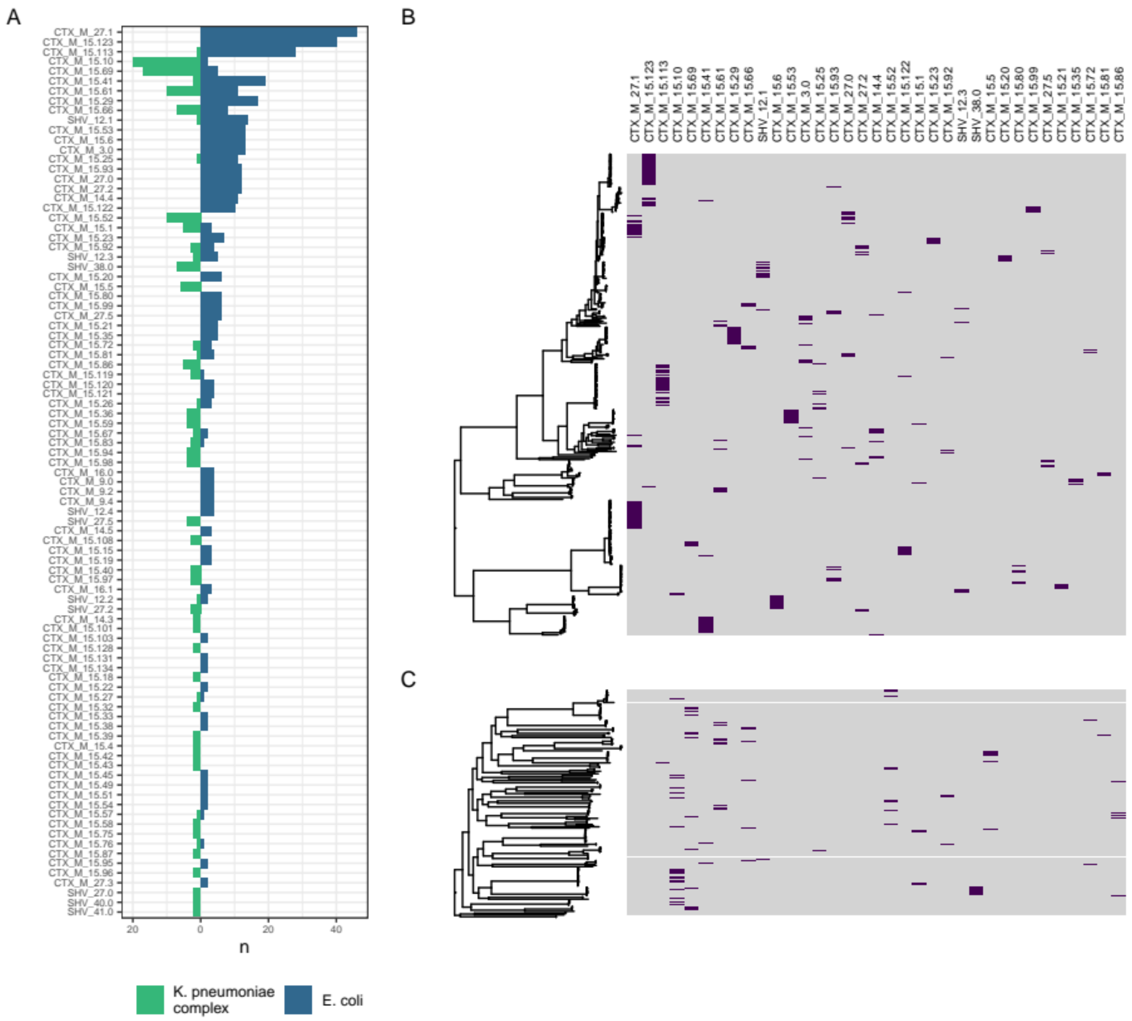


**Supplementary Figure 9:** Distribution of contig-clusters between and within genera. (A) shows distribution of contig clusters by genus. (B-C) show contig-cluster presence (purple)-absence (grey) mapped back to core gene maximum likelihood phylogeny for *E. coli* (B) and *K. pneumoniae* subsp. *pneumoniae* (C)

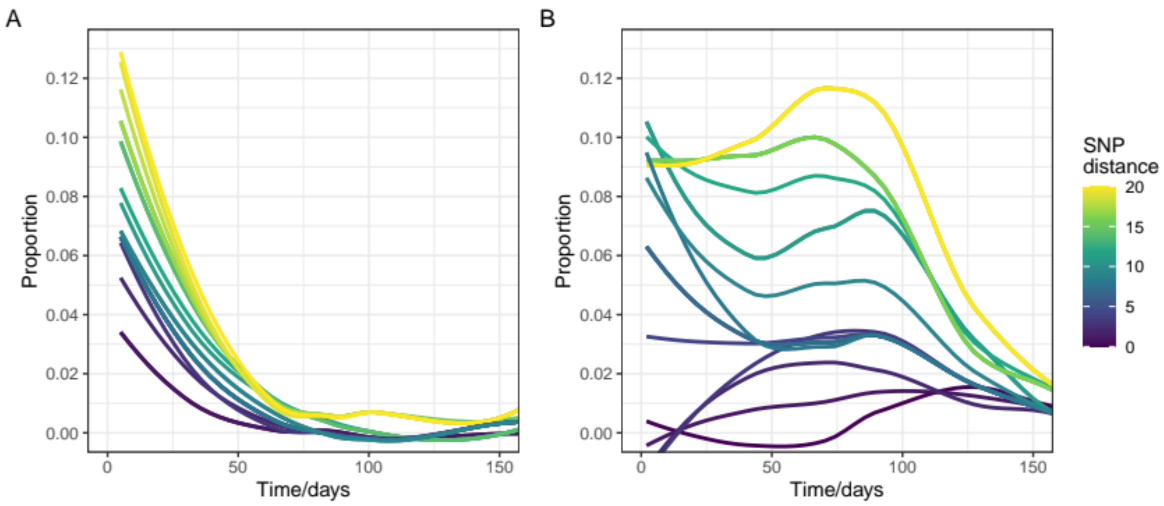


**Supplementary Figure 10:** Sensitivity analysis examining effect of changing definition of SNP-cluster from 0 to 20 SNPs. Plots show the proportion of participants who are colonised with *E. coli* (A) or *K. pneumoniae* (B) at time t = 0 who are colonised with a bacterium from the same SNP cluster (and therefore possibly the same clone) and time *t* days later

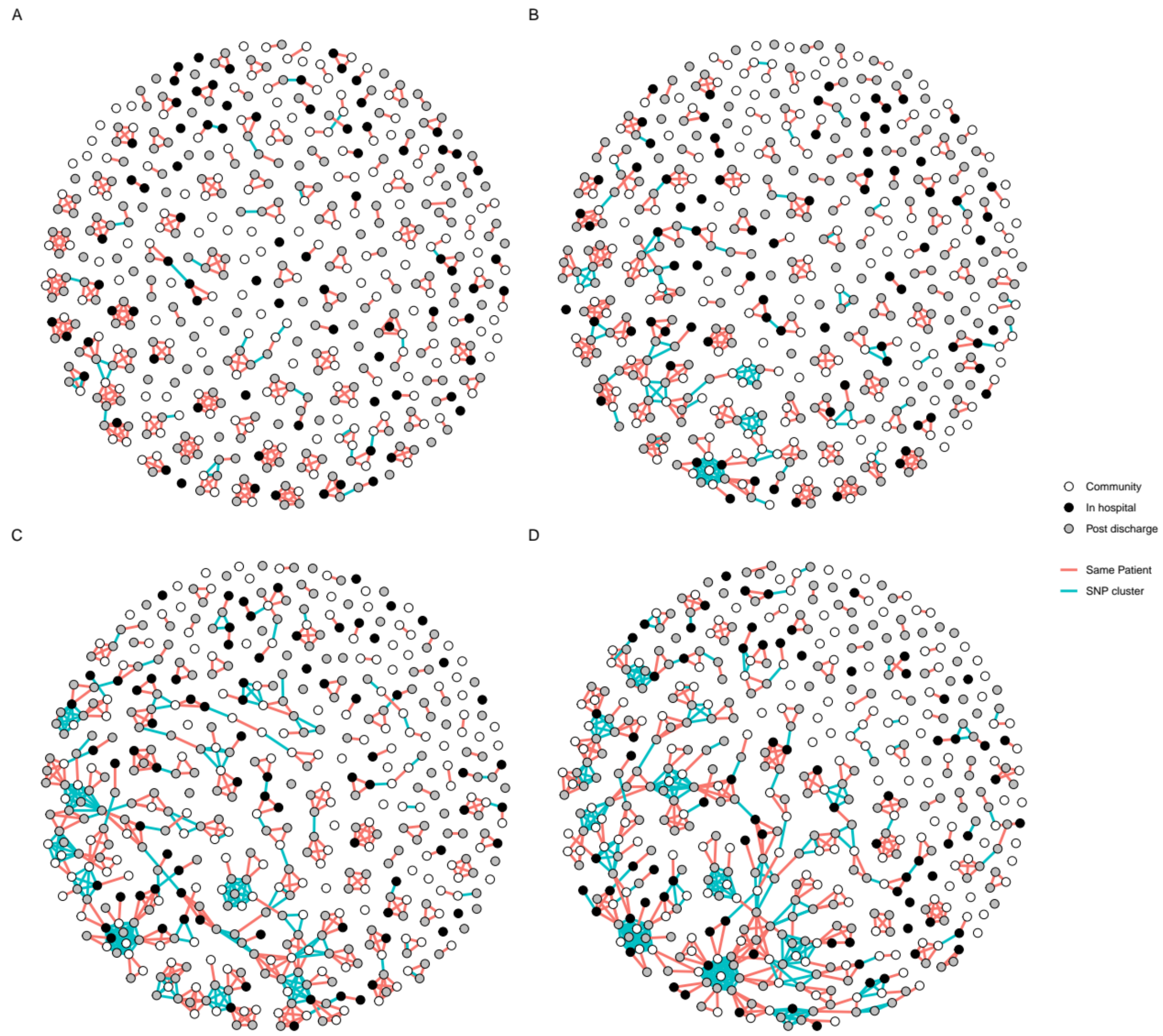


**Supplementary Figure 11:** Sensitivity analysis showing changing network plot for *E. coli* as the definition of SNP cluster is changed from 0 (A) to 3 (B), 7 (C) or 10 (D). Points are samples, coloured by place of isolation (in-hospital [black], community [white], or up to 120 days post-discharge [grey]). Red lines link samples that are within a single participant. Blue lines link samples that are differ by five or fewer SNPs.

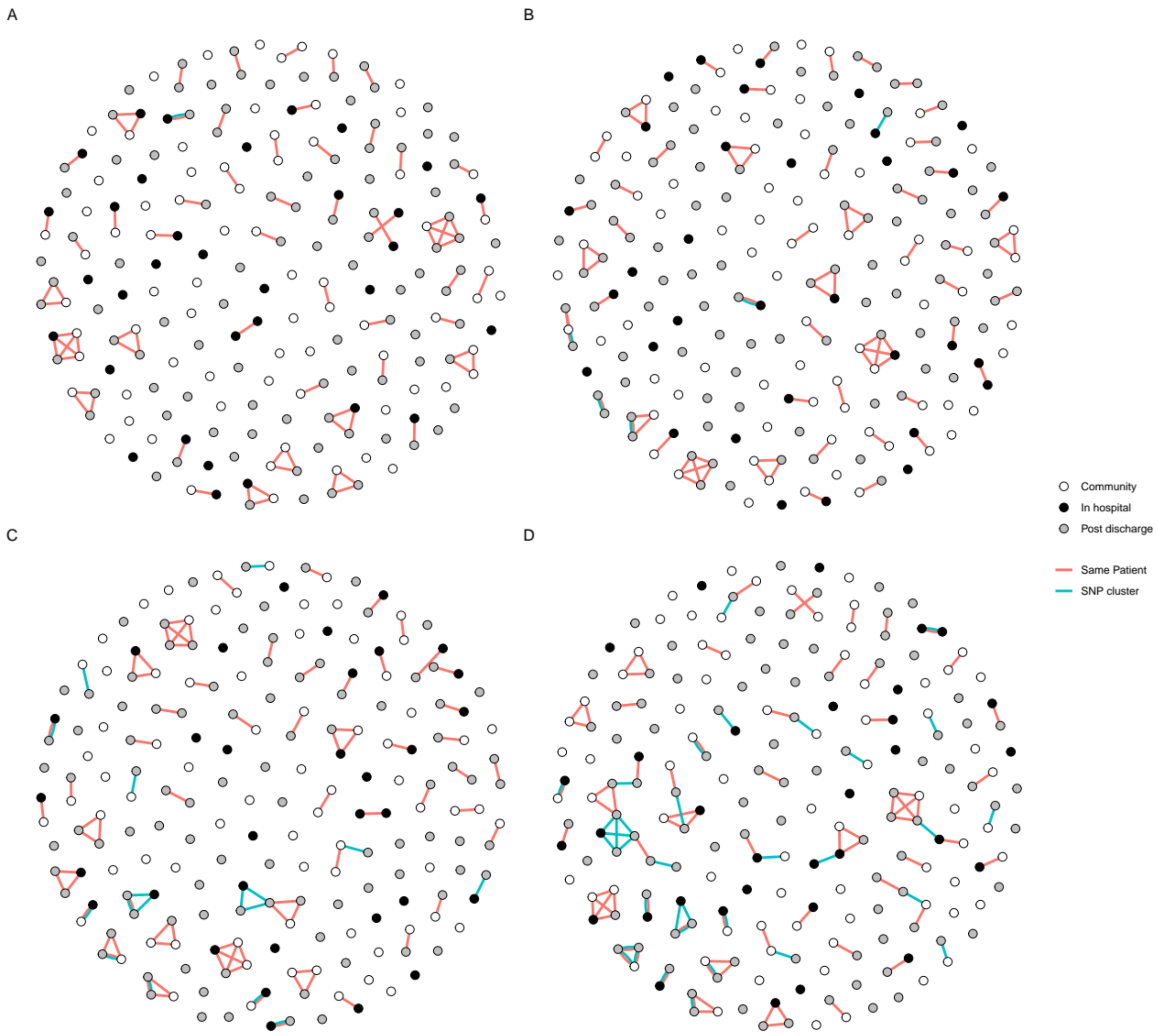


**Supplementary Figure 12:** Sensitivity analysis showing changing network plot for *K. pneumoniae* as the definition of SNP cluster is changed from 0 (A) to 3 (B), 7 (C) or 10 (D). Points are samples, coloured by place of isolation (in-hospital [black], community [white], or up to 120 days post-discharge [grey]). Red lines link samples that are within a single participant. Blue lines link samples that are differ by five or fewer SNPs.

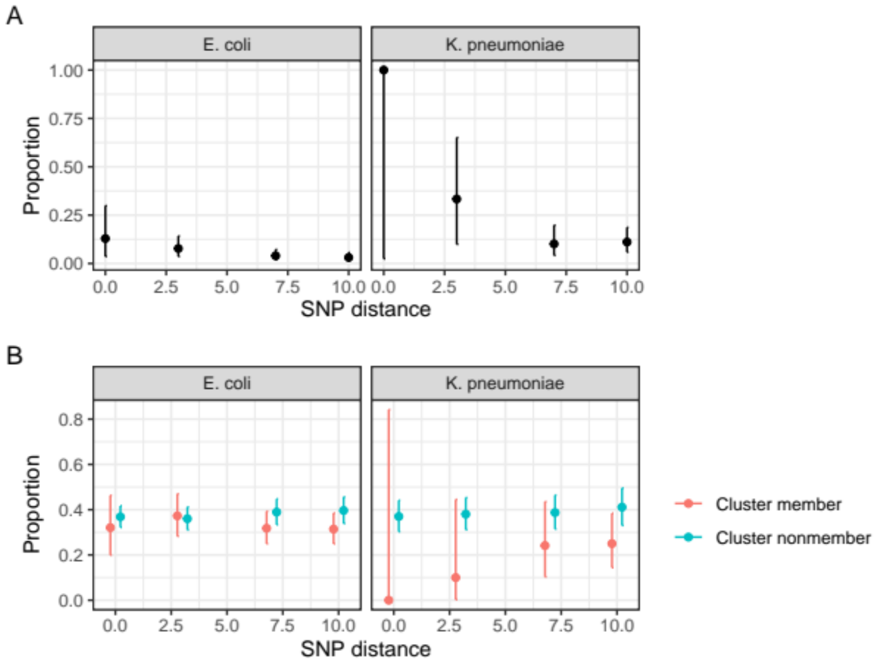


**Supplementary Figure 13:** Sensitivity analysis examining epidemiology of SNP-clusters (i.e. putative transmission clusters) as the SNP-threshold varies from 0 to 10. (A) shows the proportion of pairwise comparisons of within-SNP-cluster isolates that are within a single participant; most putative transmission clusters are between-, rather than within- participants. (B) shows the proportion of isolates that are community associated stratified by whether they are members of a putative transmission cluster or not; the proportion is similar at all threshold values, which is not suggestive of hospital-associated transmission.

**Supplementary Table 1:** ESBL carriage prevalence. *K. pneumo* here includes all *K. pneumoniae* complex isolates

| | Sepsis | | | | Inpatient | | | | Community | | | |
|---|---|---|---|---|---|---|---|---|---|---|---|---|
| **Visit** | **n** | **ESBL** | ***E. coli*** | ***K. pneumo*** | **n** | **ESBL** | ***E. coli*** | ***K. pneumo*** | **n** | **ESBL** | ***E. coli*** | ***K. pneumo*** |
| Baseline | 222 | 109 (49%) | 98 (44%) | 34 (15%) | 99 | 41 (41%) | 35 (35%) | 10 (10%) | 99 | 28 (28%) | 25 (25%) | 6 (6%) |
| Day 7 | 162 | 127 (78%) | 122 (75%) | 45 (28%) | 63 | 32 (51%) | 27 (43%) | 8 (13%) | - | - | - | - |
| Day 28 | 148 | 106 (72%) | 97 (66%) | 46 (31%) | 71 | 37 (52%) | 30 (42%) | 16 (23%) | 92 | 29 (32%) | 23 (25%) | 9 (10%) |
| Day 90 | 126 | 71 (56%) | 62 (49%) | 24 (19%) | 60 | 29 (48%) | 25 (42%) | 4 (7%) | - | - | - | - |
| Day 180 | 127 | 61 (48%) | 55 (43%) | 24 (19%) | 65 | 29 (45%) | 23 (35%) | 5 (8%) | 82 | 23 (28%) | 18 (22%) | 2 (2%) |

**Supplementary Table 2:** Antimicrobial and hospital exposure stratified by arm

| | Number exposed | | | Exposure (person-days) | | | Median (IQR) exposure length (days) | | |
|---|---|---|---|---|---|---|---|---|---|
| **Exposure** | **Sepsis** | **Inpatient** | **Community** | **Sepsis** | **Inpatient** | **Community** | **Sepsis** | **Inpatient** | **Community** |
| Total at risk | 225 | 100 | 100 | 33797 | 14336 | 21983 | 183 (63-203) | 182 (97-187) | 200 (185-219) |
| **Exposures** | | | | | | | | | |
| Hospitalised | 225 | 100 | 1 | 1727 | 500 | 1 | 5 (2-10) | 2 (2-7) | 1 (1-1) |
| Ceftriaxone | 183 | 7 | 0 | 997 | 26 | 0 | 5 (3-7) | 3 (2-4) | - |
| Co-trimoxazole | 110 | 6 | 7 | 14447 | 549 | 1388 | 180 (27-190) | 86 (6-177) | 190 (183-206) |
| Ciprofloxacin | 61 | 2 | 0 | 398 | 12 | 0 | 7 (5-7) | 6 (6-6) | - |
| TB therapy | 52 | 2 | 0 | 6843 | 291 | 0 | 178 (58-180) | 146 (133-158) | - |
| Amoxicillin | 38 | 3 | 1 | 235 | 21 | 5 | 7 (5-7) | 5 (5-8) | 5 (5-5) |
| Fluconazole | 27 | 0 | 0 | 118 | 0 | 0 | 3 (2-5) | - | - |
| Metronidazole | 24 | 2 | 0 | 148 | 10 | 0 | 6 (2-7) | 5 (5-5) | - |
| Artesunate | 11 | 0 | 0 | 25 | 0 | 0 | 2 (2-3) | - | - |
| Co-amoxiclav | 10 | 2 | 0 | 40 | 12 | 0 | 5 (2-5) | 6 (6-6) | - |
| LA | 7 | 0 | 0 | 19 | 0 | 0 | 3 (2-3) | - | - |
| Doxycycline | 7 | 0 | 0 | 34 | 0 | 0 | 3 (2-6) | - | - |
| Erythromycin | 5 | 0 | 0 | 38 | 0 | 0 | 7 (5-11) | - | - |
| Gentamicin | 4 | 0 | 0 | 15 | 0 | 0 | 4 (3-5) | - | - |
| Streptomycin | 2 | 0 | 0 | 16 | 0 | 0 | 8 (7-9) | - | - |
| Penicillin | 2 | 0 | 0 | 5 | 0 | 0 | 2 (2-3) | - | - |
| Flucloxacillin | 2 | 0 | 0 | 5 | 0 | 0 | 2 (2-3) | - | - |
| Azithromycin | 2 | 2 | 0 | 7 | 12 | 0 | 4 (3-4) | 6 (6-6) | - |
| Amphotericin | 2 | 0 | 0 | 8 | 0 | 0 | 4 (4-4) | - | - |
| Aciclovir | 2 | 0 | 0 | 47 | 0 | 0 | 24 (16-31) | - | - |
| Quinine | 1 | 0 | 0 | 1 | 0 | 0 | 1 (1-1) | - | - |

TB = tuberculosis, LA = Lumefantrine artemether. Median exposure length includes only those exposed. Total at risk shows the total number of participants and participant-days of follow up included in the study.

**Supplementary Table 3:** parameter estimates from final model

| Variable | Value |
|---|---|
| **Effect of Antibacterials** | |
| Hazard ratio ESBL-E Loss | 0.16 (0.05-0.58) |
| Hazard ratio ESBL-E Gain | 0.57 (0.16-2.25) |
| Half life of effect (days) | 43.7 (15.4-97.7) |
| **Effect of Hospitalisation** | |
| Hazard ratio ESBL-E Loss | 10.01 (1.24-52.34) |
| Hazard ratio ESBL-E Gain | 27.82 (3.60-143.18) |
| **Mean time in state** | |
| Colonised (days) | 9.7 (4.2-25.1) |
| Uncolonised (days) | 5.8 (2.5-14.3) |

In the mathematical notation used in the methods section hazard ratios are the exponential of the parameters $\alpha$ and $\beta$ in the model; half life is equal to $\gamma$log2); mean time in state assumes all other covariates are equal to zero and is then the reciprocal of $\lambda$ or $\mu$.
